# Mapping of cis-regulatory variants by differential allelic expression analysis identifies candidate causal variants and target genes of 41 breast cancer risk loci

**DOI:** 10.1101/2022.03.08.22271889

**Authors:** Joana M. Xavier, Ramiro Magno, Roslin Russell, Bernardo P. de Almeida, Ana Jacinta-Fernandes, André Duarte, Mark Dunning, Shamith Samarajiwa, Martin O’Reilly, António M. Maia, Cátia L. Rocha, Nordiana Rosli, Bruce A. J. Ponder, Ana Teresa Maia

## Abstract

Genome-wide association studies (GWAS) have identified hundreds of risk loci for breast cancer, but identifying causal variants and candidate target genes remains challenging. Since most risk loci fall in active gene regulatory regions, we developed a novel approach to identify variants with greater regulatory potential in the disease’s tissue of origin. Using genome-wide differential allelic expression (DAE) analysis on microarray data from 64 normal breast tissue samples, we mapped over 54K variants associated with DAE (daeQTLs). We then intersected these with GWAS data to reveal candidate risk regulatory variants and analyzed their cis-acting regulatory potential. We found 122 daeQTLs in 41 loci in active regulatory regions that are in strong linkage disequilibrium with risk-associated variants (risk-daeQTLs). We also identified 65 new candidate target genes in 29 of these loci for which no previous candidates existed. As validation, we identified and functionally characterized five candidate causal variants at the 5q14.1 risk locus targeting the *ATG10* and *ATP6AP1L* genes, likely acting via modulation of alternative transcription and transcription factor binding. Our study demonstrates the power of DAE analysis and daeQTL mapping to understand breast cancer genetic risk, including in complex genetic regulatory landscapes. It additionally provides a genome-wide resource of variants associated with DAE for future functional studies.

## INTRODUCTION

Genome-wide association studies (GWAS) for breast cancer (BC) have identified hundreds of risk-associated loci and have generated long lists of candidate loci requiring further validation (Wendt and Margolin, 2019). Nevertheless, the identification of the causal variants and their target genes, as well as understanding the underlying biological mechanisms, remain challenging. This is because disease risk loci often have many variants in high linkage disequilibrium (LD) with the risk-associated variant, harbor multiple genes, and mainly fall in noncoding genome regions (Gallagher and Chen-Plotkin, 2018). However, the overrepresentation of potential causal variants at active gene regulatory regions (Fachal et al., 2020; Maurano et al., 2012) indicates that variants regulating gene expression levels likely influence BC genetic predisposition, both proximally and over a long range (Darabi et al., 2015; Dunning et al., 2016; Ghoussaini et al., 2016; Meyer et al., 2011, 2008; Michailidou et al., 2017; Udler et al., 2010). These variants have commonly been mapped by expression quantitative trait loci (eQTL) analysis, but this approach is impacted by the effects of negative feedback control and environmental factors (Pastinen et al., 2006). An increasingly popular alternative approach is to detect imbalances in allelic transcript levels - differential allelic expression (DAE). By comparing the relative expression of the two alleles in a heterozygous individual, each allele will serve as an internal standard for the other, thus controlling for trans-regulatory and environmental factors affecting both alleles (Forton et al., 2007; Ge et al., 2009). Consequently, this directly indicates regulatory variants acting in *cis* - cis-acting regulatory SNPs or rSNPs.

Given the importance of cis-regulatory variants for BC susceptibility, a genome-wide map of cis-regulatory variants would be key to interpreting GWAS results and identifying causal variants of risk. Studies in various healthy tissues showed that DAE is a relatively common event (Aguet et al., 2017; Bjornsson et al., 2008; Gao et al., 2012; Ge et al., 2009; Przytycki and Singh, 2020; Romanel et al., 2015). Given that gene expression regulation is tissue-specific, performing these studies in the tissue from which the disease arises, namely, normal breast tissue, is essential. Although others have used allelic expression analysis to identify BC risk, this was carried out in tumor tissue or lymphoblastoid cells (Hamdi et al., 2014; Zhang et al., 2018). This study proposes an integrative approach to identify causal variants of risk that have a cis-regulatory role (Figure 1): to combine GWAS results with SNPs associated with DAE levels in normal breast tissue. Hence, we first carried out DAE analysis in normal breast tissue samples at a genome-wide level, then mapped the candidate risk regulatory variants for GWAS loci, and finally functionally unveiled the mechanisms underlying BC risk at a selected locus.

**Figure 1.**
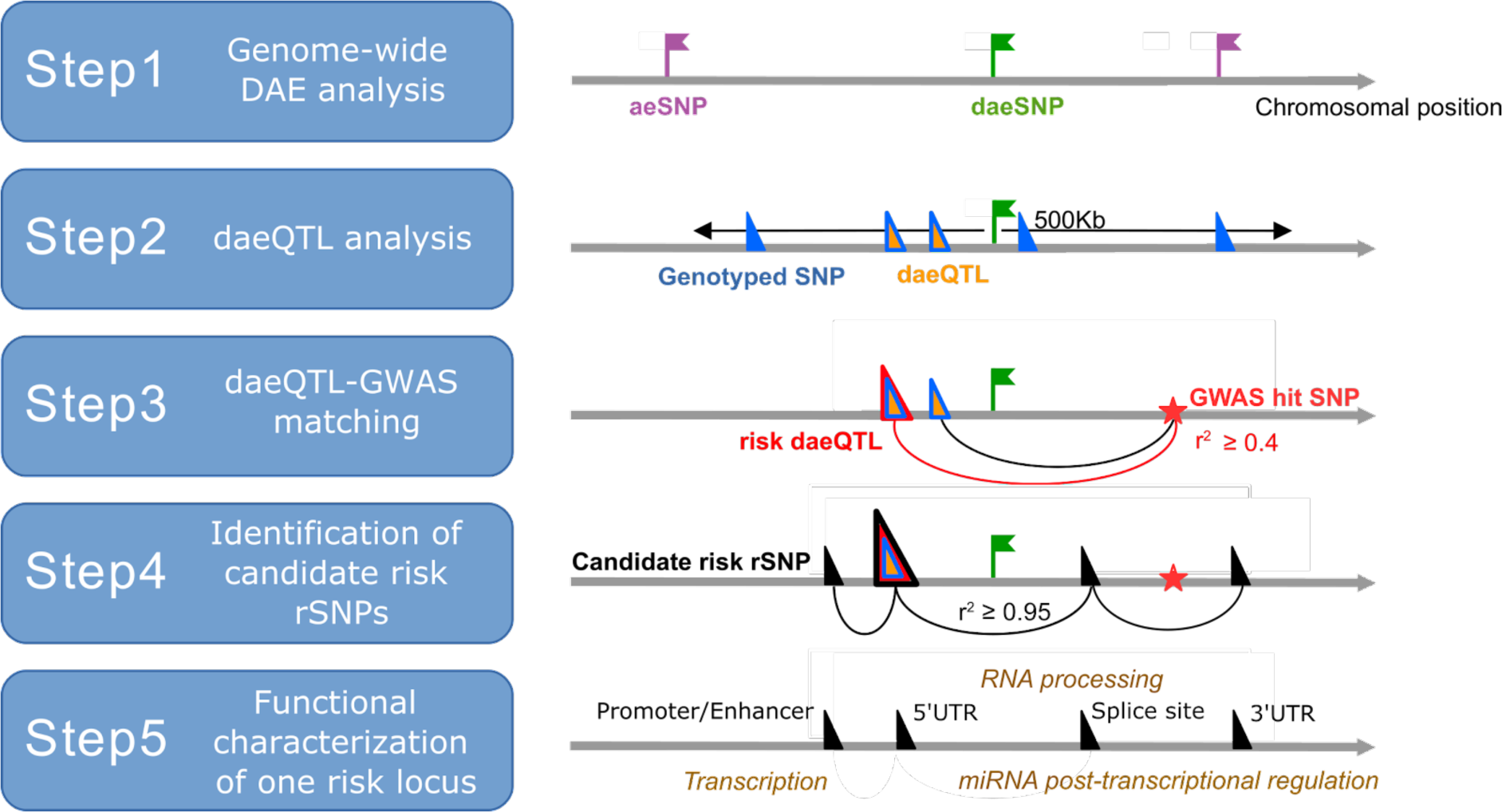
Strategy framework used to identify causal variants and target genes associated with breast cancer risk. Legend: *aeSNP* an SNP that passed quality control and at which allelic expression (AE) was measured; daeSNP an aeSNP showing differential AE (DAE); Genotyped SNP an SNP with genotype information (either genotyped in the study or imputed) and tested for association with AE ratios; daeQTL an SNP associated with AE ratios measured for a daeSNP; risk-daeQTL a daeQTL with a r^2^ ≥ 0.4 with a GWAS hit variant; candidate risk rSNP a variant with a r^2^ ≥ 0.95 with the risk-daeQTL.

## MATERIAL AND METHODS

### SNP and call filtering at the gDNA and cDNA levels

We used an Illumina Infinium Exon510S-Duo arrays dataset of normal breast tissue available from Gene Expression Omnibus (GEO, www.ncbi.nlm.nih.gov/geo/) under accession number GSE35023 (Liu et al., 2012). It consists of 66 samples of DNA and cDNA (derived from total RNA) run on Illumina Infinium Exon510S-Duo arrays. These exon-centric microarrays contain probes for 511,354 SNPs, with more than 60% of the markers located within 10 kb of a gene and targeting more than 99.9% of human RefSeq genes. Sample filtering and normalization were performed as described previously, and 12 samples were removed from further analysis (Liu et al., 2012). We performed extensive quality control after microarray data normalization and before allelic expression analysis. First, a minimum cut-off of average log_2_ RNA intensity values ≥ 9.5 for each probe was applied to remove non-expressed SNPs. Then, SNPs for which the RNA log_2_ ratios could not discriminate between homozygous and heterozygous genotypes (t Test > 0.05) were eliminated. Third, to guarantee high-quality genotyping data, a minimum call rate ≥ 90%, a Hardy-Weinberg equilibrium p-value > 1.0E-05, and at least five heterozygotes were requested for each SNP. Finally, we only kept SNPs uniquely mapped in the genome, not flagged as suspected in dbSNP149 GRCh38p7 and located in autosomes.

### Genome-wide DAE analysis

Allelic expression was measured in the filtered dataset of SNPs and samples in a varying number of individuals heterozygous (AB) for each transcribed SNP (aeSNP). As cDNA was prepared from total RNA, without selection for poly-A mRNAs, AE was measured for variants in fully processed and unspliced primary transcripts. Allelic expression ratios (AE ratios) were defined as the log_2_ of the ratio between the levels of allele A transcript and the levels of allele B transcript (heterozygote ratio), normalized by the same heterozygote ratio calculated for genomic DNA (gDNA) (Figure S1), to account for copy number variation and correct for technical biases. Differential allelic expression (DAE) was called at the sample level when AE ratios were greater than 0.58 or less than -0.58 (corresponding to the log_2_ of a 1.5-fold difference between alleles).

aeSNPs were classified as monoallelically expressed SNPs (maeSNPs) (Gimelbrant et al., 2007) when presenting more extreme AE ratios and a random distribution of heterozygotes above 0.58 and below -0.58 ratios, suggesting the expression of only one allele with a random choice between alleles. Genes with at least one maeSNP were labeled maeGenes.

After filtering out maeSNPs, the remaining aeSNPs were tested with the Equal or Given Proportions test (prop.test function in R), with the alternative hypothesis that the proportion of heterozygotes with absolute AE ratios ≥ 0.58 is greater than 10% for any given SNP. The resulting p-values were corrected using a false discovery rate of 5% to define daeSNPs (Figure S1, Figure 1 – Step 1). Genes with at least one daeSNP were henceforth denominated daeGenes.

Validation of nine daeSNPs was performed by TaqMan^®^ PCR technology, as described previously (Maia et al., 2009), in 25 independent normal breast tissue samples heterozygous for a variable number of individuals per SNP using the following TaqMan® Genotyping Assays predesigned by Applied Biosystems: C 8354687_10; C 29939330_20; C 31232634_10; C 3133316_10; C 11844169_10; C 2627792_10; C 1517694_1_; C 787630_20; C 3108259_10. The prop.test was equally applied to confirm the presence of differential allelic expression.

### Annotation of variants

Variants were annotated according to hg38/GRCh38 with biomaRt v 2.40.5. aeSNP consequence types were categorized as follows: UTR if classified as 3_prime_UTR_variant or 5_prime_UTR_variant; coding if classified as coding_sequence_variant, incomplete_terminal_codon_variant, missense_variant, stop_retained_variant, synonymous_variant, stop_lost, start_lost, stop_gained, splice_region_variant, splice_acceptor_variant or splice_donor_variant; intronic if classified as intron_variant; and noncoding_transcript_variant if classified as noncoding_transcript_variant, noncoding_transcript_exon_variant or mature_miRNA_variant. We classified aeSNPs further according to gene biotype as follows: pseudogene if located in IG_C_pseudogene, processed_pseudogene, transcribed_unprocessed_pseudogene, transcribed_unitary_pseudogene, translated_unprocessed_pseudogene, unprocessed_pseudogene, unitary_pseudogene, transcribed_processed_pseudogene, polymorphic_pseudogene or rRNA_pseudogene; protein-coding gene if located in protein_coding, IG_V_gene, TR_C_gene, TR_J_gene, TR_V_gene or TEC; and noncoding_rna if located in lncRNA, miRNA, misc_RNA, snRNA, snoRNA, scaRNA or ribozyme.

To test whether classes of consequence type and gene biotype were overrepresented (i.e., enriched) in the list of daeSNPs, we applied a one-tailed Fisher’s exact test (alternative = “greater”). Information from imprinted genes was retrieved from a comprehensive study of genomic imprinting in the breast (Goovaerts et al., 2018) and from the geneimprint database (http://www.geneimprint.com) searching for Imprinted Genes: by Species: Human.

### Genotype imputation

Imputation was run on the Illumina Exon 510 Duo germline genotype data from the 64 samples that passed microarray quality control filters. Before imputation data, quality control was applied to the genotyping data, and SNPs with call rates < 85%, minor allele frequency < 0.01, and Hardy-Weinberg equilibrium with p-value < 1.0E-05 were excluded from the analysis. Imputation was performed using MACH1.0 (Li et al., 2010) and the phased haplotypes for HapMap3 release (HapMap3 NCBI Build 36, CEU panel - Utah residents with Northern and Western European ancestry) as a reference panel. We applied the recommended two-step imputation process: model parameters (crossover and error rates) were estimated before imputation using all haplotypes from the study subjects and running 100 iterations of the Hidden Markov Model (HMM) with the command option - greedy and -r 100. Genotype imputation was then carried out using the model parameter estimates from the previous round with command options of -greedy, -mle, and -mldetails specified. Imputation results were assessed by the platform-specific measures of imputation uncertainty for each SNP (rq Score) and filtered for an rq-score≥0.3, as suggested in the author webpage (http://csg.sph.umich.edu/abecasis/mach/tour/) and MAF ≥ 0.01.

### Candidate rSNP mapping

Mapping of candidate rSNPs associated with the DAE observed - henceforth designated as daeQTLs (differential allelic expression quantitative trait loci) (Figure S1, Figure 1 - Step 2) - took into consideration the pattern of AE ratio distribution displayed by each daeSNP, as this is highly dependent on the LD between the daeSNP and the rSNP acting upon the gene (Xiao and Scott, 2011).

To test the association between candidate SNP zygosity and the allelic expression of a daeSNP, henceforth designated daeQTL analysis (differential allelic expression quantitative trait loci analysis), we considered the pattern of the allelic expression (AE) ratio distribution displayed at each daeSNP, as this is dependent on the linkage disequilibrium between the daeSNP and the rSNP. When a single rSNP is in strong LD (r^2^ ∼1) with the daeSNP, the normalized AE ratios for all heterozygotes will be unidirectional, with all samples preferentially expressing the same allele (i.e., all samples exhibiting either positive AE ratios or negative AE ratios). In this case, a one-sample Wilcox test was used to compare the mean normalized AE ratios for samples heterozygous for the candidate rSNP to 0. When the rSNP is not in r^2^ ∼1 with the daeSNP, the distribution of the AE ratios will depend on the rSNP-daeSNP haplotypes present in the analyzed samples, and we applied a two-sample Wilcox test for the null hypothesis that the absolute AE ratios at the samples heterozygous for the candidate rSNP are higher than the absolute AE ratios at the samples homozygous for the tested rSNP. These tests were performed for rSNP-daeSNP pairs located within 500 kb of each other. P-values were adjusted with the Benjamini‒Hochberg method (Hochberg and Benjamini, 1990), using all daeSNP/tested SNP pairs, with the distance between them as a covariate (package ihw, R) (Ignatiadis et al., 2016) and reported as significant when the false discovery rate was below 5%.

### Breast cancer GWAS data retrieval

Nine hundred and sixty-eight GWAS-significant risk-associated SNPs for BC published until April 2018 were retrieved from the NHGRI-EBI Catalog of published genome-wide association studies (GWAS Catalog) (MacArthur et al., 2017) using the gwasrapidd R package (Magno and Maia, 2019). Filters included a significance level cut-off p-value ≤ 1.0E-05 and the reported traits: “Breast cancer”, “Breast cancer (early onset)”, “Breast cancer (estrogen-receptor negative)”, “Breast cancer (male)”, “Breast cancer in BRCA1 mutation carriers”, “Breast cancer in BRCA2 mutation carriers”, “Breast cancer male”, and “Breast cancer and/or colorectal cancer”. The complete list of SNPs is presented in Table S1.

### Proxy SNP retrieval

Variants in LD with index SNPs were retrieved from Ensembl (Yates et al., 2015) using the function get_ld_variants_by_window() from the ensemblr R package (https://github.com/ramiromagno/ensemblr) using the 1000 GENOMES project data (phase_3) for the CEU population and a genomic window size of 500 kb (250 kb upstream and downstream of the queried variant). The r^2^ cut-off used varied between 0.2 and 0.95 depending on the analysis and is indicated in each analysis description.

### Retrieval of previously suggested BC target genes

Genes previously suggested as targets of cis-acting regulatory variation in post-GWAS studies for BC, with extensive fine-scale mapping and *in silico* prediction or functional analysis, and those classified as *Inquisit 1* by Fachal and colleagues (Fachal et al., 2020) are indicated in Table S2.

### GTEx eQTL and gene expression data retrieval

The Genotype-Tissue Expression (GTEx) project identified expression quantitative trait loci (eQTL) using normal mammary tissue samples (Consortium et al., 2015). eGenes (genes with at least one SNP in cis significantly associated, at a false discovery rate (FDR) of ≤0.05, with expression differences of that gene) and significant variant-gene associations based on permutations were downloaded from GTEx Analysis V8 (dbGaP Accession phs000424.v8.p2, available on 18/07/2019).

All SNP-gene associations tested for breast mammary tissue, including nonsignificant and gene expression levels (TPM), were downloaded from GTEx Analysis V7 (available on 2016-01-15).

### Comparison of daeGenes, eGenes and gwasGenes

First, the list of publicly available eGenes was compared with the daeGenes identified in our study, restricting this comparison to genes analyzed in both datasets. Then, we investigated the percentage of gwasGenes, defined as genes containing variants in moderate to strong LD (r^2^≥0.4) with GWAS index SNPs, displaying evidence of cis-regulation by either DAE or eQTL analysis.

### Functional characterization of candidate risk SNPs

Candidate risk rSNPs were examined for regulatory potential by assessing the overlap of the variant’s location with epigenetic marks derived from the ENCODE (Dunham et al., 2012) and NIH Roadmap Epigenomics project data (Kundaje et al., 2015) using the R package haploR. Candidate causal variants (variants overlapping with DNase I hypersensitivity sites and H3K4me1 or H3K4me3 or histone modifications in normal breast or breast tumor cell lines) at the 5q14.1-14.2 locus were further analyzed regarding their genomic context and transcription factor (TF) binding using the UCSC Genome Browser (Gonzalez et al., 2021; Kent et al., 2002), HaploReg v4.1 (Ward and Kellis, 2012) and RegulomeDB v1.1 (Boyle et al., 2012) tools. Emphasis was given to overlapping with transcription factor (TF) binding identified in breast myoepithelial cells (BR. MYO, E027), human mammary epithelial cells (HMECs, E119), variant human mammary epithelial cells (vHMECs, E028) and two BC cell lines (MCF-7 and T47D). Allele-specific epigenetic modifications (H3k4me3 and DNase I), RNA polymerase II (POL2), and transcription factors (TF) binding with alignment data available in HMEC, MCF-7 and MCF-10A breast cancer cell lines from ENCODE were retrieved and visualized with the Integrative Genomics Viewer (IGV Version 2.3.71) tool (Thorvaldsdóttir et al., 2013), to analyze protein‒DNA interactions and allelic preferential binding. Differential allelic binding was analyzed in heterozygous candidate risk rSNPs located within TF binding peaks in experiments with a read coverage at the SNP site ≥ 20. We applied a two-tailed binomial test with the null hypothesis assuming no bias (balanced binding of the protein to the two alleles of the variant). The p-value was corrected for multiple testing using the R package qvalue (Storey et al., 2021). When multiple tracks for the same SNP, trait, and cell line existed, only the p-value for the experiment with higher total read counts was reported in the main manuscript.

Analysis related to alternative transcription at the 5q14.1-14.2 locus was carried out in three ways. First, sQTLseekeR (v1.4) (Monlong et al., 2014) was used to test the association of genetic variants with alternative isoform expression in both normal breast and tumor tissue using total read counts derived from RNA-seq data from the TCGA (TCGA-BRCA, hg19) and GTEx (phs000424.v6.p1, hg38) projects. Only *ATG10* displayed sufficient alternative transcription dispersion to allow sQTL analysis. Additionally, all SNPs within 5 kb upstream or downstream of *ATG10* were included in the analysis, not only the candidate risk rSNPs, to increase the stringency of the association exercise. P-values for all SNPs tested for ATG10 sQTL analysis were controlled for multiple testing using a 5% FDR. Correlation analyses between -log10 (FDR q-value) and LD (r^2^) with rs7707921 were performed using Pearson’s test. Then, the overlapping of variant location with RNA processing-associated proteins was assessed using CLIP data retrieved from POSTAR2 (http://lulab.life.tsinghua.edu.cn/postar/) (Zhu et al., 2019) and from RBP-Var (http://www.rbp-var.biols.ac.cn/) (Mao et al., 2016), which additionally informed on riboSNitch potential (Corley et al., 2015). Finally, allele-specific RBP binding predictions were performed with RBPmap (Paz et al., 2014) using the analyzed variant flanking sequence (30 nucleotides on each side, with the variant at index 31) using all available human RBP motifs.

### Haplotype analysis

Haplotypes in the 5q14.1-14.2 region were analyzed on Haploview 4.2 using the imputed genotypes from the 64 normal breast tissue samples (Barrett et al., 2005). For candidate risk SNPs whose genotype was not possible to determine (because it was neither genotyped nor imputed), a proxy SNP in strong LD (r^2^ ≥ 0.95) was used instead. Haplotype blocks were generated using the default algorithm.

### TCGA-BRCA gene expression analysis

Processed gene expression and isoform expression from RNA-Seq data for 113 normal solid tissues and 1102 primary solid tumors from the TCGA-BRCA project, together with corresponding clinical data, were retrieved from the Genomic Data Commons archive using the R package TCGAbiolinks (Colaprico et al., 2016) accessed in October 2018. Isoform expression was annotated according to the genome assembly hg19, and total gene expression was annotated according to hg38. We applied two-sample Wilcoxon tests to compare the mean expression of *ATG10* isoforms between normal-solid tissues (normal-matched) and breast tumors, correcting for multiple testing with the Benjamini and Hochberg (BH) procedure. We applied Pearson’s test to correlate gene expression among *ATG10*, *RPS23*, and *ATP6AP1L*. Spearman’s test was applied to correlate *ATG10*, *RPS23*, and *ATP6AP1L* with *MYC* and *MAX* gene expression.

## RESULTS

### Cis-regulatory variation is common in normal breast tissue

Genome-wide allelic expression (AE) analysis was performed using microarray data from 64 normal breast tissue samples. Normalized allelic expression ratios were calculated for SNPs in coding and noncoding regions upon filtering for the cDNA signals’ expression level and allelic discrimination potential. Overall, we identified 91,467 autosomal allelic-expressed SNPs (aeSNPs) located in 21,527 annotated Ensembl genes (median of three aeSNPs per gene) (Figure S2). Unsurprisingly, the number of aeSNPs analyzed per gene correlated with the annotated gene length (rho = 0.60, p-value < 2.2e-16, Figure S3).

We found that almost one-third of the aeSNPs (26,266 out of 91,467) displayed biallelic differential expression (daeSNPs, q-value ≤ 0.05) (Table 1, Table S3), while 84 SNPs displayed monoallelic expression (maeSNPs). TaqMan PCR validated seven out of nine daeSNPs (Figure S4) that showed significant DAE and concordant preferential expression (Fisher’s exact test p-value > 0.05).

**Table 1.**
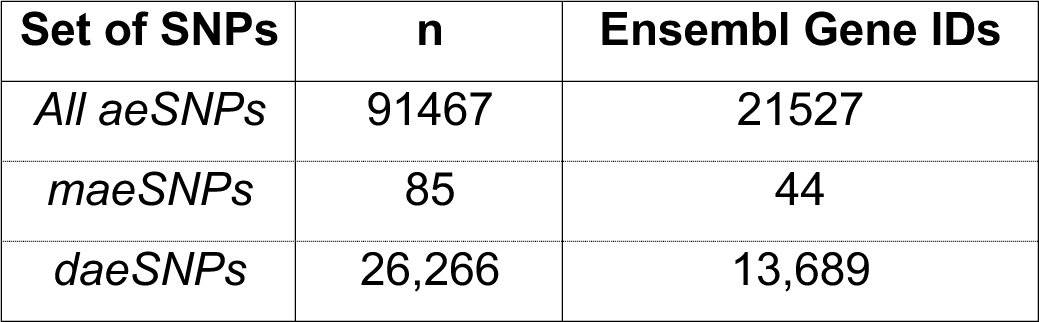
Summary of the genome-wide breast tissue allelic expression analysis results.

The daeSNPs are distributed across the genome, with low interchromosomal variability (ranging from 26 to 35%, Figure S5). They overlapped 13,688 (65%) annotated genes (daeGenes), of which 3,666 (17%) harbored three or more daeSNPs (Figure 2a, Table 1, Table S3). When considering daeSNPs mapping exclusively to one gene, we identified 8,193 daeGenes (out of 12,944) that showed evidence of being under the control of allele-specific cis-acting factors, either genetic or epigenetic. In terms of consistency of DAE detection across the length of these genes, we found that in the majority of daeGenes, the frequency of daeSNPs was higher than 40% (7476 in 13,688), with 3894 daeGenes presenting imbalances in all the analyzed aeSNPs (Figure 2b). The aeSNPs showed a large distribution of mean |AE ratios|, with daeSNPs centered at 0.60 (corresponding to a difference between alleles of 1.5) and non-daeSNPs centered at 0.26 (corresponding to a difference of 1.2). Twelve percent of daeSNPs showed average absolute AE ratios between 1 and 5, corresponding to average allelic fold changes ranging from 2 to 34 (Figure 2c, Table S3). The amplitude of the imbalances measured at aeSNPs correlated negatively with the average expression level of both alleles (rho = -0.4, p-value < 2.2e-16) (Figure 2d) but not with the standard deviation across individuals (Figure S6). The aeSNPs are located mainly in intronic regions and noncoding transcript genes, but non-daeSNPs and daeSNPs showed differences in class distribution for consequence type, with daeSNPs enriched at unannotated regions (p-value < 0.01, Figure 2e). Although most of the aeSNPs analyzed were in protein-coding genes, daeSNPs were relatively more common in noncoding genes and pseudogenes when compared to non-daeSNPs (p-value < 0.01, Figure 2f).

**Figure 2.**
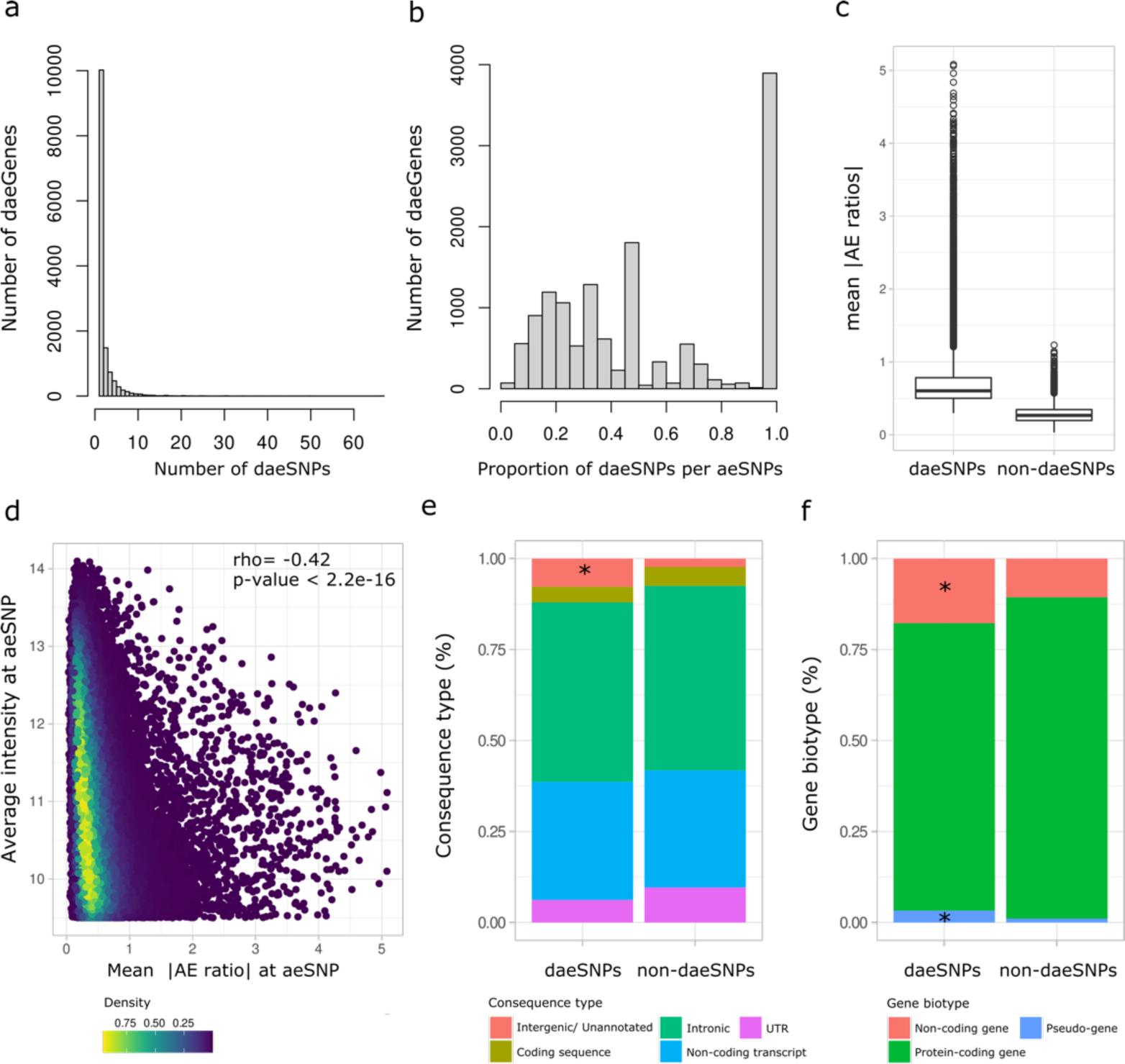
Characterization of aeSNPs. a) Histogram of the rank number of daeSNPs identified per gene across 17135 annotated genes. b) Histogram of the rank proportion of daeSNPs per aeSNPs identified per gene. c) Box plot with the distribution of the mean of the absolute values of AE ratios across heterozygous individuals measured at non-daeSNPs and daeSNPs. d) Distribution of the mean absolute values of AE ratios at aeSNPs according to the average intensity of both alleles at aeSNPs in the microarray Spearman’s results of a Spearman’s correlation test are shown. e) and f) Relative frequency of aeSNPs and daeSNPs according to consequence type and gene biotype, respectively. (*) denotes the classes for which daeSNPs were enriched (p<0.01).

### Monoallelic expression in breast tissue

Regarding monoallelic expression, maeSNPs were annotated to 44 Ensembl genes (Table 1, Table S4, Figure S7), the majority of which were previously reported as imprinted in breast tissue (e.g., *IGF2* or *ZDBF2*) or in other tissues (e.g., *KCNQ1*, *KCNQ1OT1*, *RTL1*, *NAA60*, *ZIM2,* and *L3MBTL1*), validating our AE analysis. Interestingly, we detected maeSNPs in a region containing the lncRNA *MEG9* and a cluster of miRNA genes that had only previously been reported as imprinted in nonhuman species (Hagan et al., 2009; Seitz et al., 2003; Tierling et al., 2006). Additionally, we found unreported monoallelic expression at an intergenic region (22q11.23), suggesting the existence of unannotated transcripts in this region. Notably, we observed two groups of heterozygotes preferentially expressing opposite alleles of rs17122278, an intronic variant of *ARCN1*, suggesting the latter as a candidate novel monoallelically expressed protein-coding gene in breast tissue.

### Mapping of daeQTLs in normal breast tissue

Evidence of DAE supports that a gene’s expression is controlled by cis-regulatory variation, which can be mapped using AE ratios as a quantitative trait – in what we termed DAE quantitative trait loci (daeQTL) analysis. Here, we found a minority of daeSNPs (6928 out of 26266) for which all the heterozygotes preferentially expressed the same allele. This pattern indicates moderate to strong linkage disequilibrium between the daeSNP and the rSNPs acting on it (Xiao and Scott, 2011) [27]. Hence, our mapping approach considered the allelic expression (AE) ratio distribution pattern displayed at each daeSNP, and one-sample or two-sample Wilcox tests were applied accordingly. We identified 54357 daeQTLs (5% FDR) for 6761 (26%) daeGenes (Table S5), primarily located within 20 kb from the corresponding daeSNP but as far as the 500 kb window used for the analysis (Figure 3a). daeQTLs for *MROH8 and ZNF132,* two coding genes located on chromosomes 19 and 20, respectively, were among the most significant ones found, but we identified other highly significant daeQTLs (adjusted p-values smaller than 5.0E-04) for 2507 genes.

**Figure 3.**
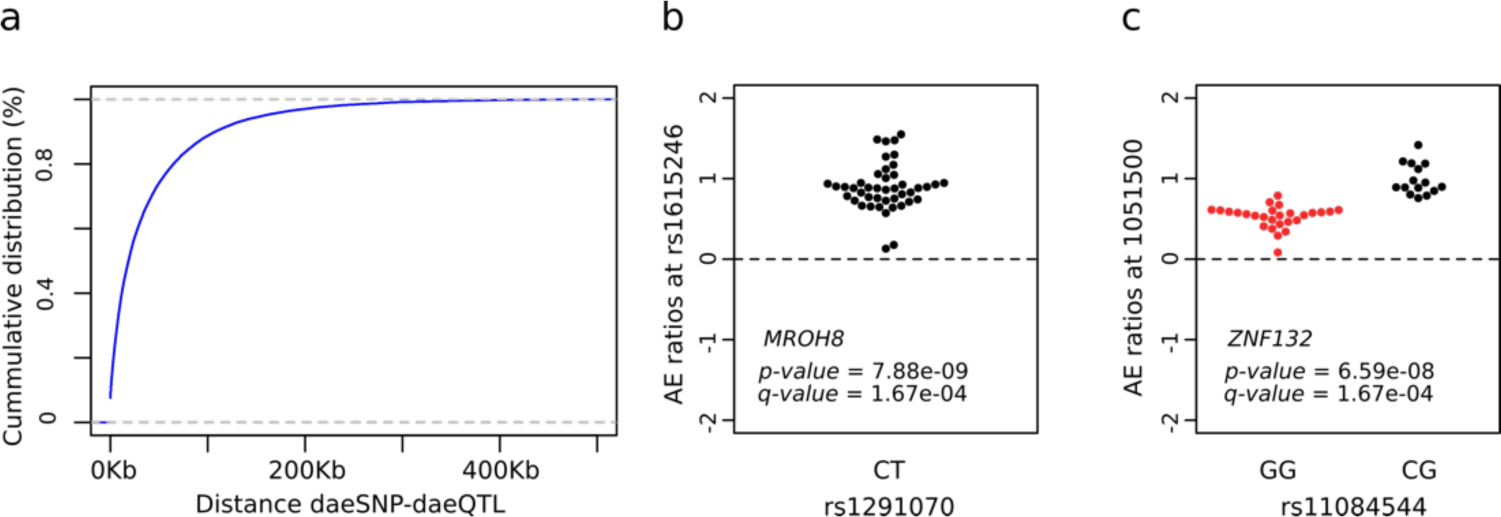
Mapping of variants associated with differential allelic expression. a) Empirical cumulative distribution for the distance between the daeSNP and corresponding mapped daeQTL. b) and c) daeQTL mapping result for the most significant daeQTL identified for *MROH8* using a one-sample Wilcox test and for *ZNF132* using a two-sample Wilcox test. The AE ratios calculated at the daeSNPs are represented on the y-axis in the two panels and stratified according to genotype at the candidate SNP (black dots represent heterozygous individuals, and red dots represent homozygous individuals).

### Identification of target genes within BC risk loci

To pinpoint the most likely candidate target genes within BC risk loci, a main post-GWAS challenge, we identified the genes within previously reported GWAS loci (gwasGenes) displaying the most robust evidence of being under the control of cis-regulatory variation, provided either by DAE (daeGenes) or eQTL (eGenes) analysis. We found that most gwasGenes (783 out of 948) showed such evidence, with 69% of these with evidence via DAE analysis (358 genes identified solely by DAE and 300 by both analyses) (Table S6). Compared to all genes studied, gwasGenes presented a significant enrichment of Fisher’s significance of DAE (Fisher’s exact test = 2.48e-05). Finally, we successfully mapped daeQTLs for 385 gwasGenes (Figure 4, Table S6).

**Figure 4.**
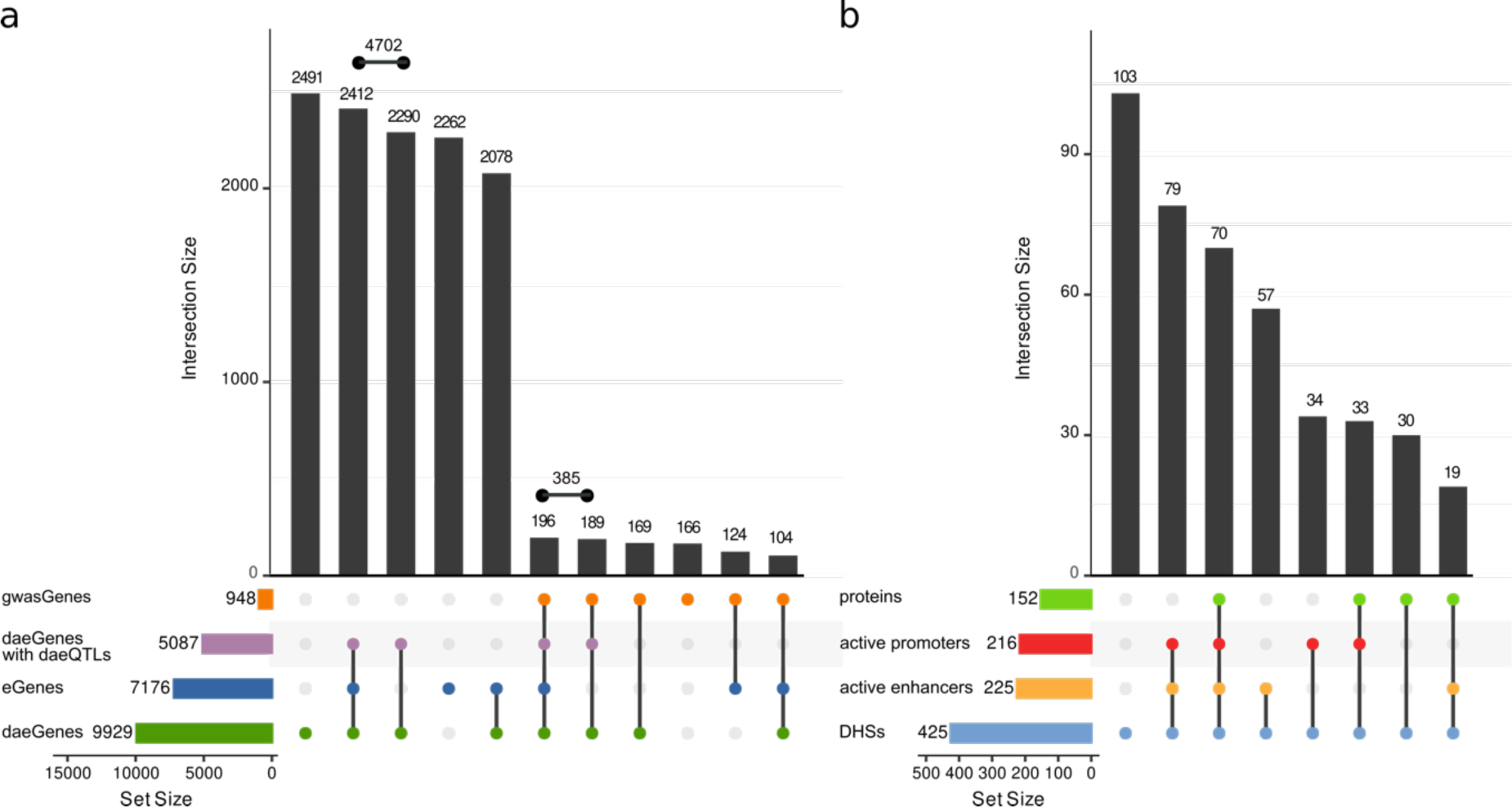
Summary of colocalization analyses for daeGenes and risk-daeQTLs. **a)** UpSet plot for 15,706 genes tested for DAE and eQTL (GTEx breast mammary tissue). Legend: daeGenes genes identified as having differential allelic expression in normal breast tissue; eGenes genes reported as being eQTL genes in GTEx mammary tissue data (q-value ≤ 0.05); gwasGenes genes where GWAS index SNPs or proxies (r^2^ ≥ 0.4) are located; daeGenes with daeQTL mapping daeGenes for which daeQTLs were identified. b) UpSet plot for 425 variants located in DHSs, according to the presence of protein binding and location in active promoters and/or enhancers in breast cell lines.

Next, we verified our ability to identify 178 previously proposed breast cancer target genes (Table S2). We found that 44% of these genes were exclusively daeGenes (e.g., *ELL, TOX3,* RNF115), 23% were both daeGenes and eGenes (e.g., *CASP8, POU5F1B, STXBP4*) and 14% were exclusively eGenes (e.g., *RMND1, HELQ, PRKRIP1*). However, we did not find evidence supporting other genes, such as C*ITED4, IGFBP5, and MYC.* (Table S2). As total levels of gene expression may confound the ability to identify daeGenes and eGenes, it is noteworthy that eGenes showed higher median levels overall than daeGenes and only 4.7% of exclusive daeGenes showed low median levels (<0.1 TPM) (Figure S8).

### Identification of causal variants within BC risk loci

Another post-GWAS challenge we addressed was the identification of the causal variants within risk loci. We first identified 1431 daeQTLs in moderate to strong LD (r^2^ ≥ 0.4) with GWAS index SNPs (Figure 1 – Step 3) (GWAS p-value < 1.0E-05), henceforth referred to as risk-daeQTLs. These were distributed across 93 loci in 19 chromosomes, primarily in introns, followed by intergenic regions (Table S7, Figure S9). Then, we assessed these risk-daeQTLs plus their proxies (r^2^ ≥0.95) for their cis-acting regulatory potential. We started by identifying 425 variants located in DNase I hypersensitivity sites (DHS), of which the majority (69%) mapped to regions with histone marks associated with active regulatory elements (Figure 4b, Figure S10). More specifically, 149 risk-daeQTLs co-localized with both active promoter (H3K4me3 and H3K9ac) and active enhancer-associated (H3K4me1 and H3K27ac) histone marks, 76 co-localized exclusively with active enhancer-associated marks and another 67 exclusively co-localized with active promoter-associated marks. Of these, 122 risk-daeQTLs also showed protein binding evidence, thus representing strong candidate causal variants within 41 of the initial 93 BC risk loci (Table S8).

Among these 41 risk loci, we detected 47 novel candidate target genes in 29 loci with no previous report of target genes, such as *SMC2* in 9q31.1, *MLLT10* in 10p12.32, and *MAN2C1* and *PTPN9* in 15q24.2. We confirmed previously reported target genes in nine loci and identified eight novel genes, including *NASP* and *IPP* in 1p34.1 and *ATP6AP1L* in 5q14.1. Finally, we identified strong candidate causal variants at six loci but could not discern the target gene due to a lack of genomic annotation (Table 2).

**Table 2.**
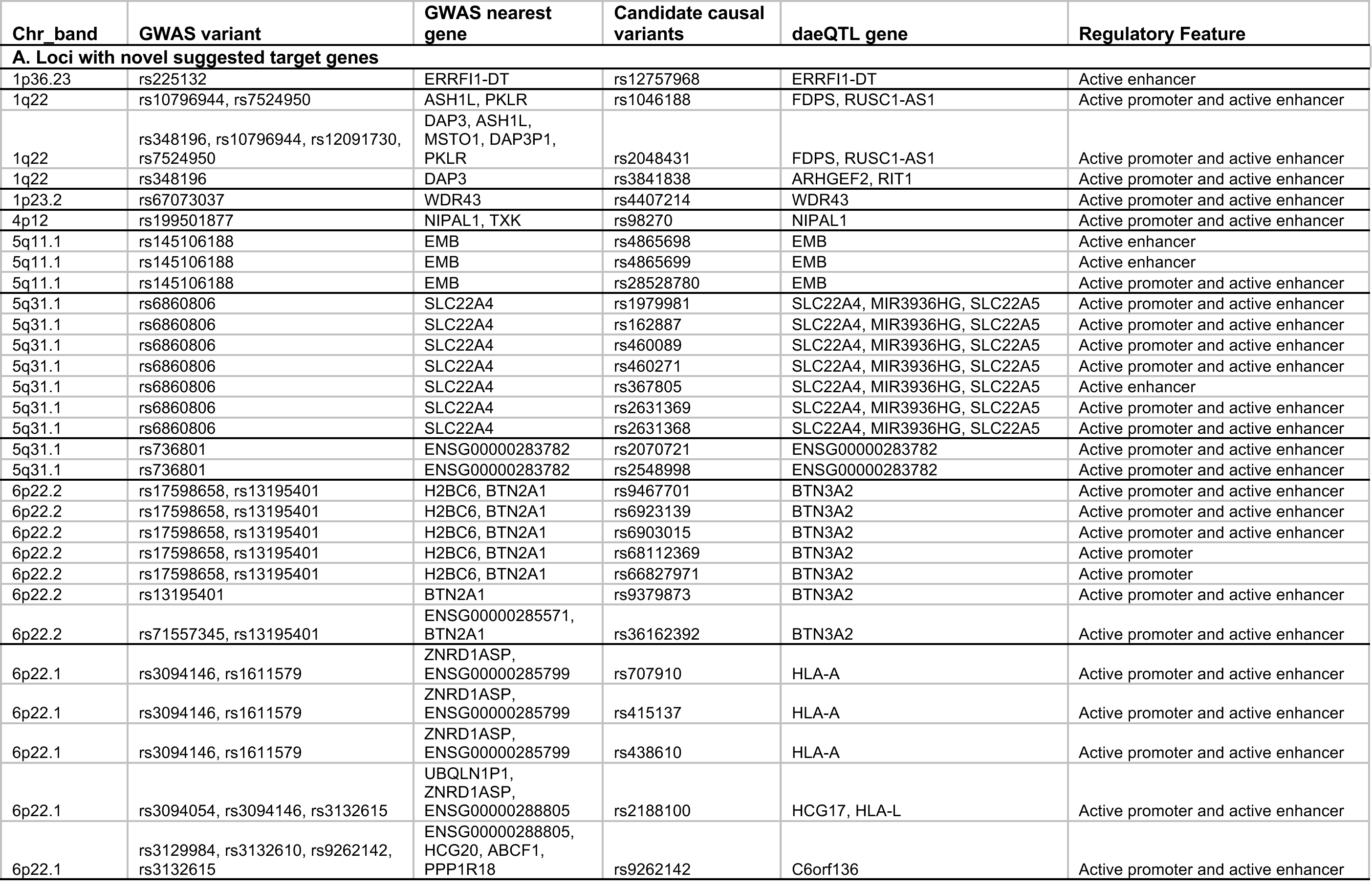
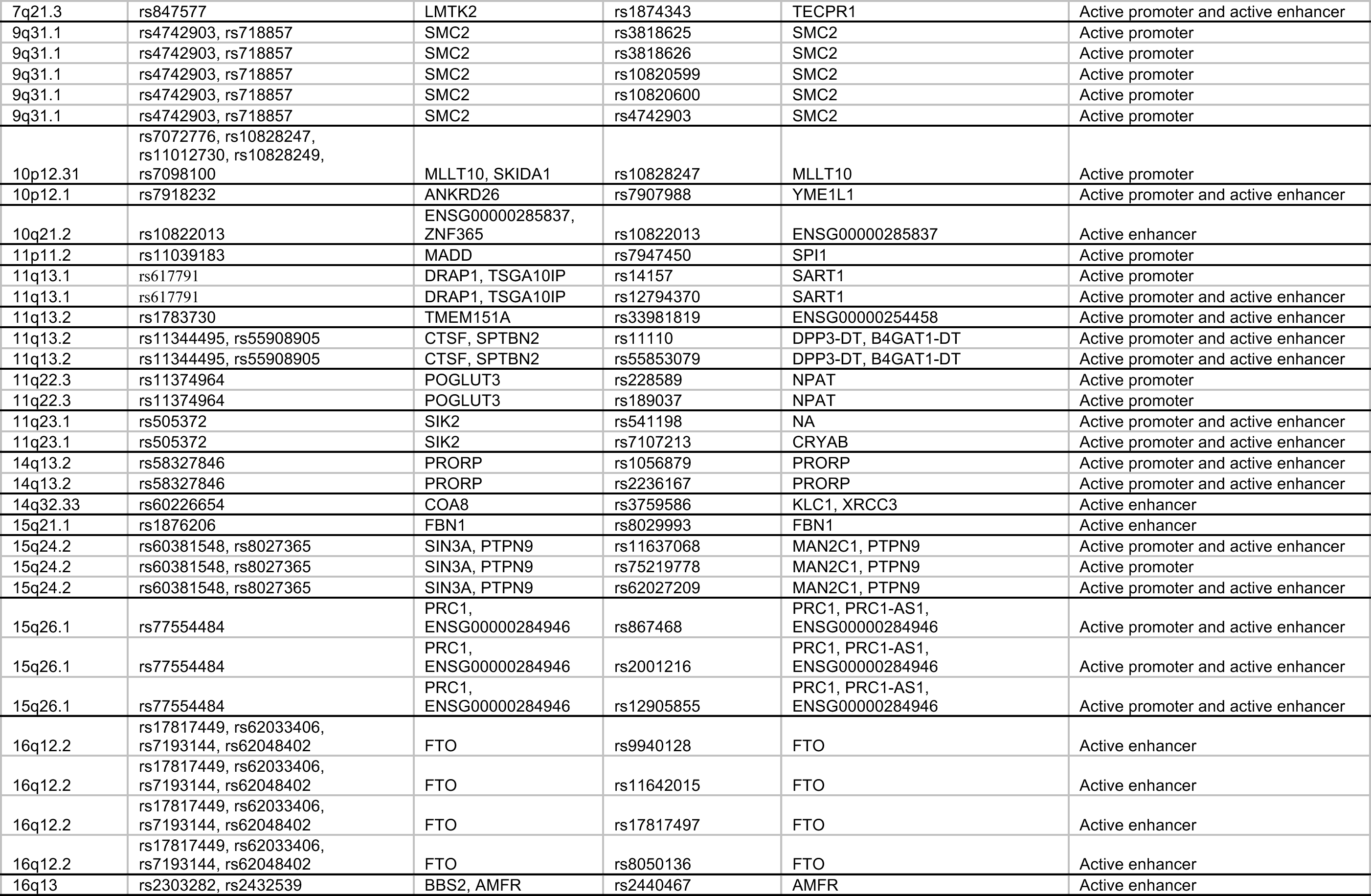
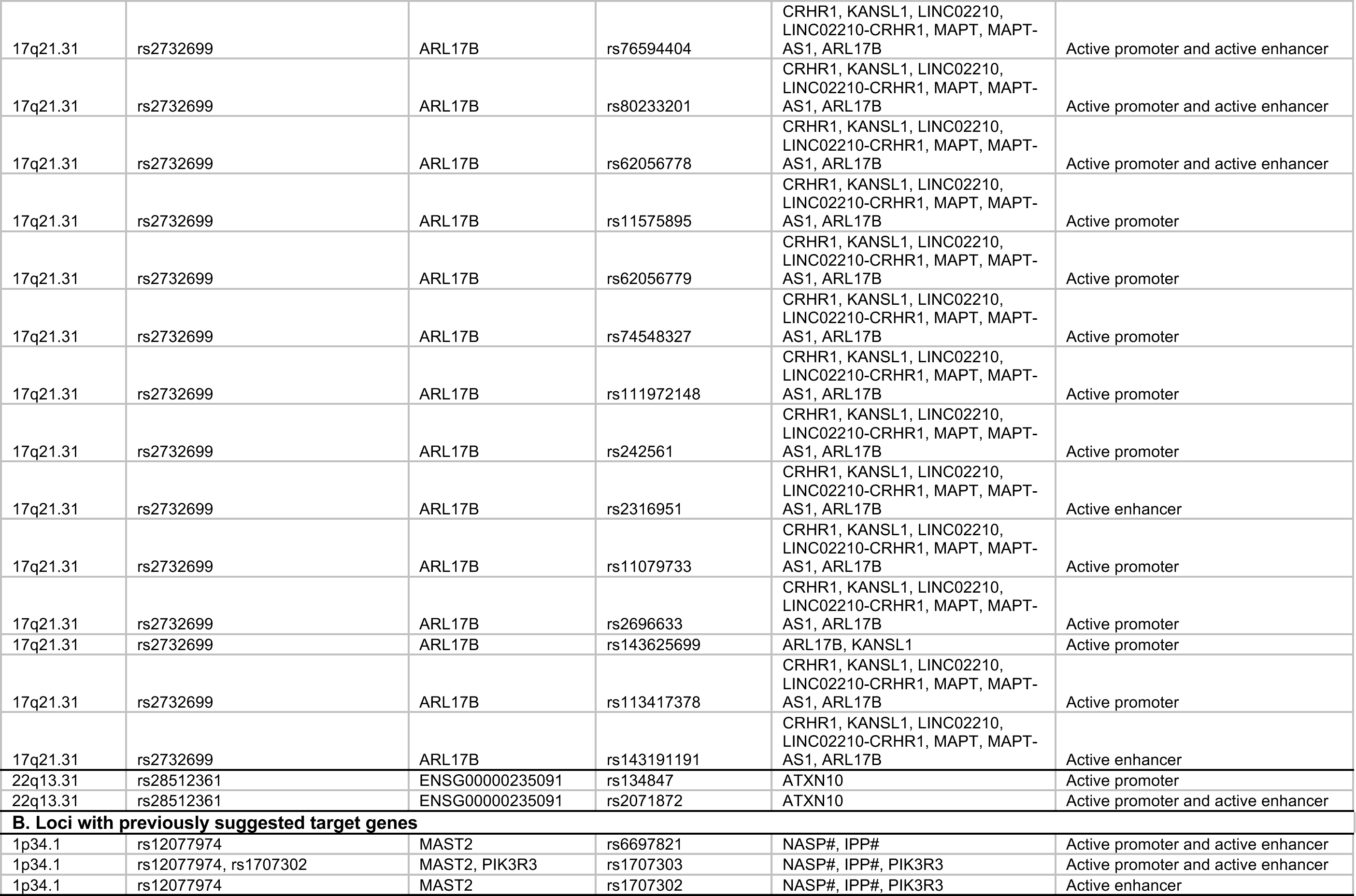
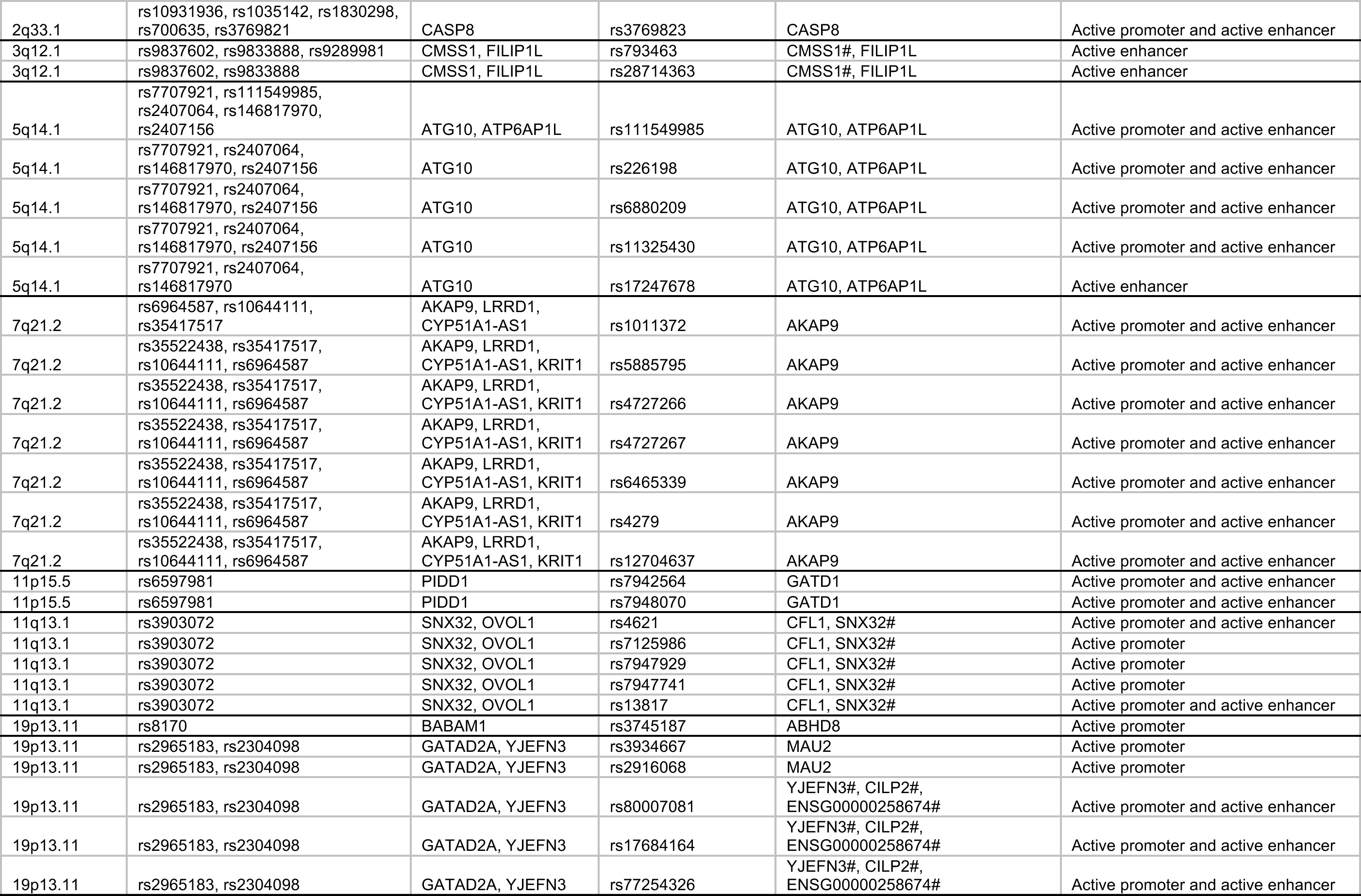
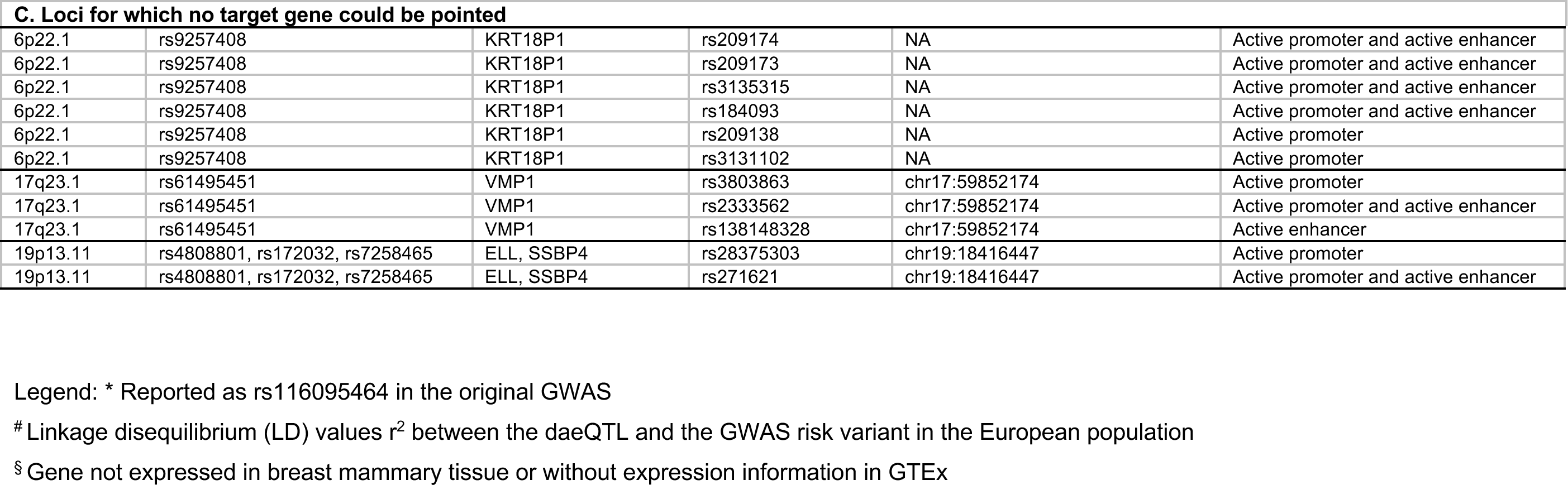
Loci with candidate risk rSNPs and novel suggested target genes.

Notably, 2222 daeQTLs were also in lower LD with GWAS hits (0.2 ≤ r^2^ <0.4), representing a valuable dataset warranting further exploration (Table S9).

### Mapping of cis-regulatory risk variants at the 5q14.1-14.2 locus

To further show the potential use of our integrated approach, we focused our follow-up studies on the BC risk locus 5q14.1-14.2, where some of the most significant risk-daeQTLs and candidate causal variants were identified. In this locus, rs7707921 was previously associated with BC risk in two meta-analyses (OR for alternative A allele = 1.07, 95% CI = [1.05-1.1], p=5E-11) (Michailidou et al., 2017, 2015). The region containing this intronic variant of *ATG10*, its proxy variants (r^2^ ≥ 0.4), and other risk-associated variants reported in this locus spans three genes (*ATG10*, *RPS23*, and *ATP6AP1L*), hindering the identification of the causal variant(s) and their target gene(s) in this locus.

First, all three genes showed DAE, supporting their regulation by cis-regulatory variants: 10 daeSNPs out of 37 aeSNPs at *ATG10*, one daeSNP out of two aeSNPs at *RPS23,* and three daeSNPs out of five aeSNPs at *ATP6AP1L* (Figure S11). The highest mean |AE ratios| detected at daeSNPs in these genes was 1.27 (2.4-fold) at *RPS23*, followed by 0.92 (1.9-fold) at *ATP6AP1L* (Figure 5 - panel 2, Figure S11). By daeQTL mapping analysis, we identified daeQTLs for all three genes: 56 for *ATG10 (*spreading along the *ATG10-ATP6AP1L* region), 4 for *RPS23 (*limited *to RPS23-ATP6AP1L)* and 26 for *ATP6AP1L (*spreading along the *ATG10-ATP6AP1L* region) (Figure 5 - panels 3 to 5). Additionally, we classified as risk-daeQTLs the 38 *ATG10* daeQTLs and 24 *ATP6AP1L* daeQTLs (22 of which are common to the two genes) in moderate to strong LD (r^2^ ≥ 0.4) with the risk-associated variants. Furthermore, both *ATG10* and *ATP6AP1L* daeQTL analysis results strongly correlated with the corresponding LD with the GWAS lead-SNP rs7707921 (Figure S12), further supporting the role of variants regulating the expression of these two genes in the risk for breast cancer.

**Figure 5.**
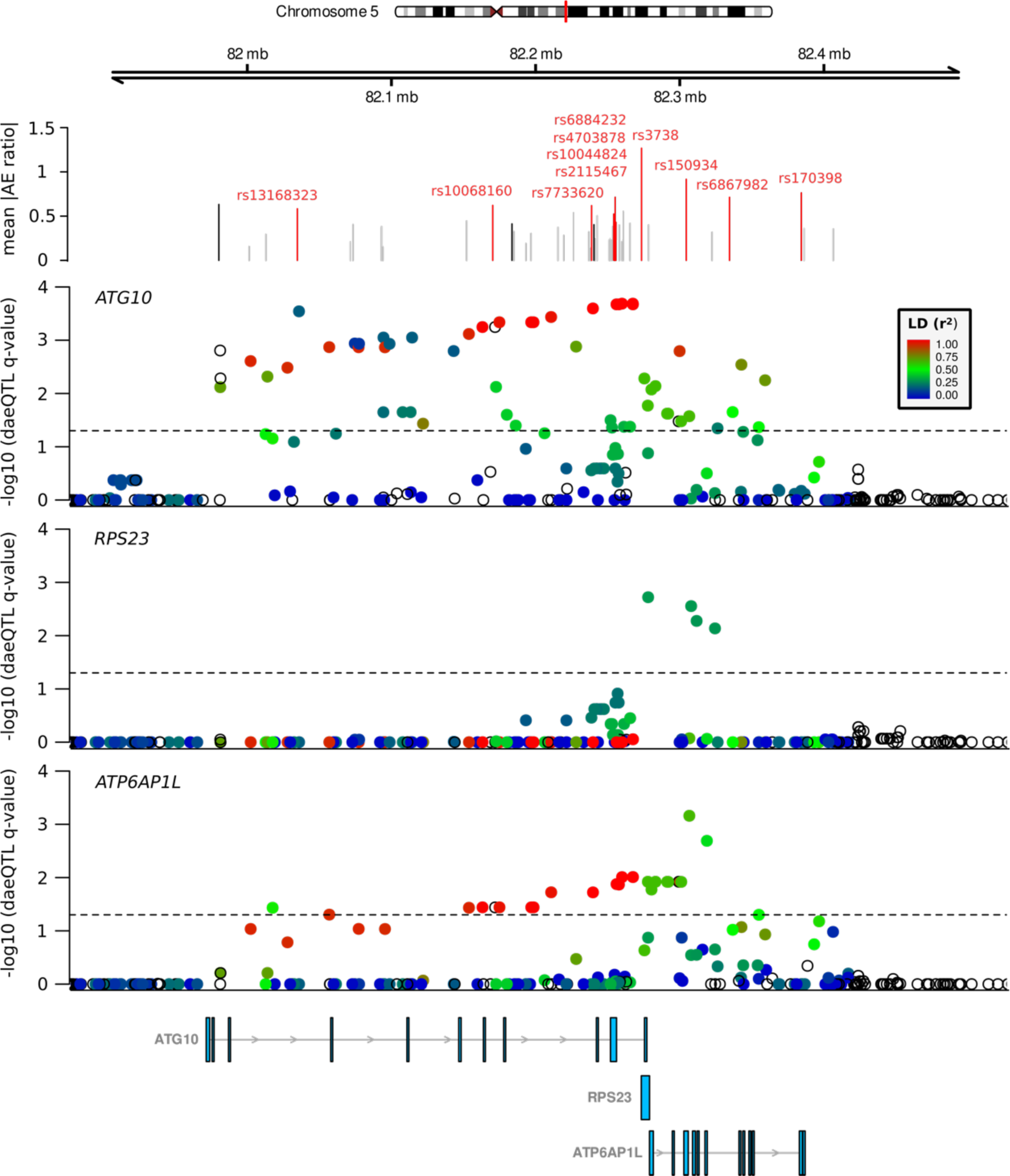
Evidence of DAE and daeQTL analysis at the 5q14.1 BC risk locus. The top track shows the mean values of the absolute AE ratios measured at aeSNPs across the region, with the non-daeSNPs shown in grey, the daeSNPs in black, and the daeSNPs with mapped daeQTLs in red. The subsequent tracks show the daeQTL mapping corrected p-values for *ATG10*, *RPS23,* and *ATP6AP1L*.

### Cis-regulatory risk variants act via two different mechanisms on genes in the 5q14.1-**14.2 locus**

The overlap analysis of the risk-daeQTLs with epigenetic marks in breast cell lines identified seven candidate causal variants for *ATG10* and *ATP6AP1L* (Table S8, Table S10). We investigated these variants further for allelic differences in transcription factor binding and association with histone modifications and DHSs. One of these SNPs, rs111549985, overlies the active promoter of *ATG10* (Figure S13), and its minor G-allele is preferentially associated with the H3K4Me3 modification in HMECs (2.7-fold, p = 3.7e-03) and shows robust preferential binding by POL2 in MCF7 cells (9-fold, *p* = 4.0E-04). However, DHS was more significantly associated with the major/reference C allele in T47D cells (0.5-fold, *p* = 4.6e-05) (Figure 6a, Table S11). Another two candidate causal variants, rs226198 (intronic to *RPS23*) and rs688025’UTR (located at *RPS23* 5’UTR) overlay the shared promoter of *RPS23* and *ATP6AP1L* and a predicted enhancer interacting with the *ATG10* promoter (Figure S14). The minor C-allele of rs226198 showed preferential binding by MYC and MAX transcription factors, which are known to cooperate in cancer (Dang, 2012) (12.6-fold and 7.9-fold difference, respectively, *p* < 2.2e-16) and preferential H3K4me3 marking (2.7-fold, *p* = 1.4e-02) in MCF-7 cells (Figure 6b, Table S11). It would be interesting to elucidate whether rs226198 impacts the binding of both factors and H3K4me3 deposition or whether this epigenetic mark is a consequence of altered transcription, as previously suggested (Floc’hlay et al., 2020; Howe et al., 2017). The minor T-allele of rs6880209 also showed preferential binding by MYC (4.8-fold, *p* < 2.2e-16) and MAX (2.4-fold, *p* = 2.7e-03), with smaller fold-change differences than rs226198, and additional preferential binding by POL2 (2.6-fold, *p* = 1.27e-06) in MCF7 cells. However, similar to rs111549985, DHS preferentially occurred in the major/reference C-allele in T47D cells (5.3-fold, *p* = 9.1e-04) (Figure 6c, Table S11). Interestingly, the expression of *MAX* correlated with *ATG10, RPS23,* and *ATP6AP1L,* and the expression of *MYC* correlated with the expression of *ATG10* (Figure S15). Furthermore, the expression levels of *ATG10* and *ATP6AP1L* were positively correlated in breast tissue from healthy women (top 2.5% quantile of 500,000 pairwise tests) and in normal-matched tissue from patients with BC (Figure S16). The observation that *ATG10* and *ATP6AP1L* are in different topologically associating domains (TADs) and that the candidate causal variants rs226198 and rs6880209 fall on the boundary between them (Figure S17) suggests that a shared pattern of chromatin condensation does not drive the correlated gene expression but instead by a shared cis-regulatory sequence.

**Figure 6.**
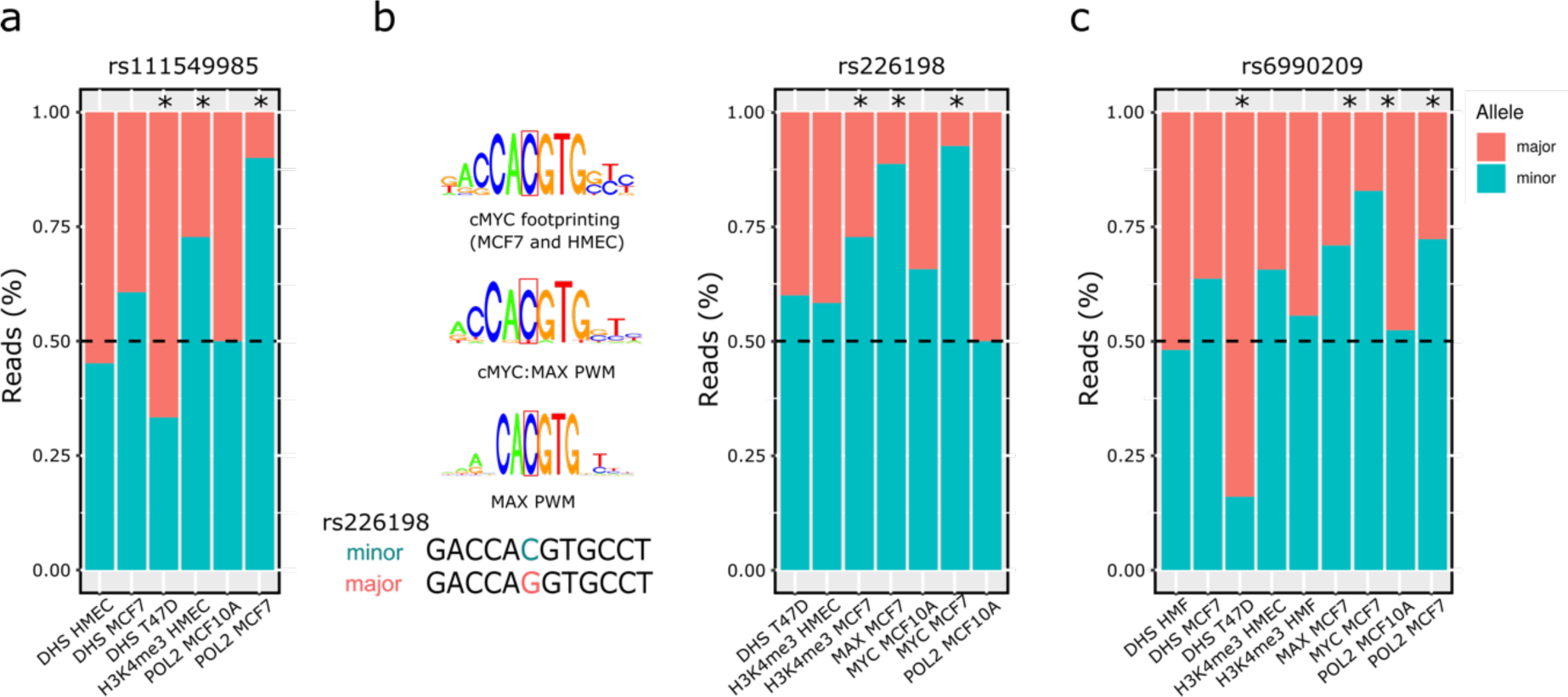
Variants at the 5q14.1 risk locus associated with differential transcription factor binding. Allele-specific analysis of the effect of three candidate risk rSNPs — (a) rs111549985, (b) rs226198, and (c) rs6880209 — on RNA polymerase II (POL2) and transcription factor (TF) binding, DNase I targeting (DHS) and H3K4me3 modification in different heterozygous cell lines. An asterisk indicates statistically significant imbalances (two-sided binomial test, p-value ≤ 0.05). Legend: HMEC human mammary epithelial cells; MCF7 human breast (adenocarcinoma) cell line; T47D human breast tumor cell line; MCF10A human breast epithelial cell line.

Since genetic variants affecting mRNA decay or alternative splicing (Robles-Espinoza et al., 2021) can cause allelic expression imbalances, we aimed to explore further the role of alternative transcription in gene expression regulation and in driving risk at the 5q14.1 locus. To accomplish this, we performed an sQTL analysis for *ATG10* that was not restricted to the candidate risk rSNPs but included all SNPs located within 5 kb upstream and downstream of *ATG10* to increase the stringency of the exercise.

We identified six sQTLs (FDR ≤ 5%) in the tumor data, whose minor alleles were associated with changes in the expression of two protein-coding isoforms: decreased expression of ENST00000458350 (one extra exon) and increased expression of ENS3’UTR0282185 (longer 3’UTR) (Figure 7, Figure S18a, Table S12). Interestingly, ENST00000282185 is expressed at significantly lower levels in tumors than in normal-matched tissue, in line with the reported oncogenic effect of UTR length (Mayr and Bartel, 2009), although with a small effect size (fold-change = 1.20) (Figure S19). The strong correlation between sQTL q-values and LD with the lead GWAS SNP rs7707921 (r=0.94, p-value = 3.15E-12, Figure S17b) supports the contribution of alternative transcription of *ATG10* to BC risk. Although no sQTL was detected for *ATG10* in normal breast data (Table S12), sQTL nominal p-values and LD with rs7707921 were still correlated in normal matched breast samples (r = 0.59, p-value = 0.002) (Figure S20). *ATP6AP1L* did not display sufficient alternative transcription dispersion to allow the sQTL analysis. Subsequent functional analysis of ATG10’s sQTLs, and their proxy SNPs (LD r2 ≥ 0.95), revealed the prediction of rs111549985 (5’UTR) and rs6884232 (3’UTR) to cause a riboSNitch (a functional RNA structure disrupted by an SNP (Corley et al., 2015)). Although RBP binding data for breast tissue do not exist, these variants have been reported to disrupt the binding of Xrn2 (involved in termination by RNA polymerase II) and of Igf2bp1 (a translation regulator) in K562 cells (Table S13, Table S14), which would require confirmation in breast cells.

**Figure 7.**
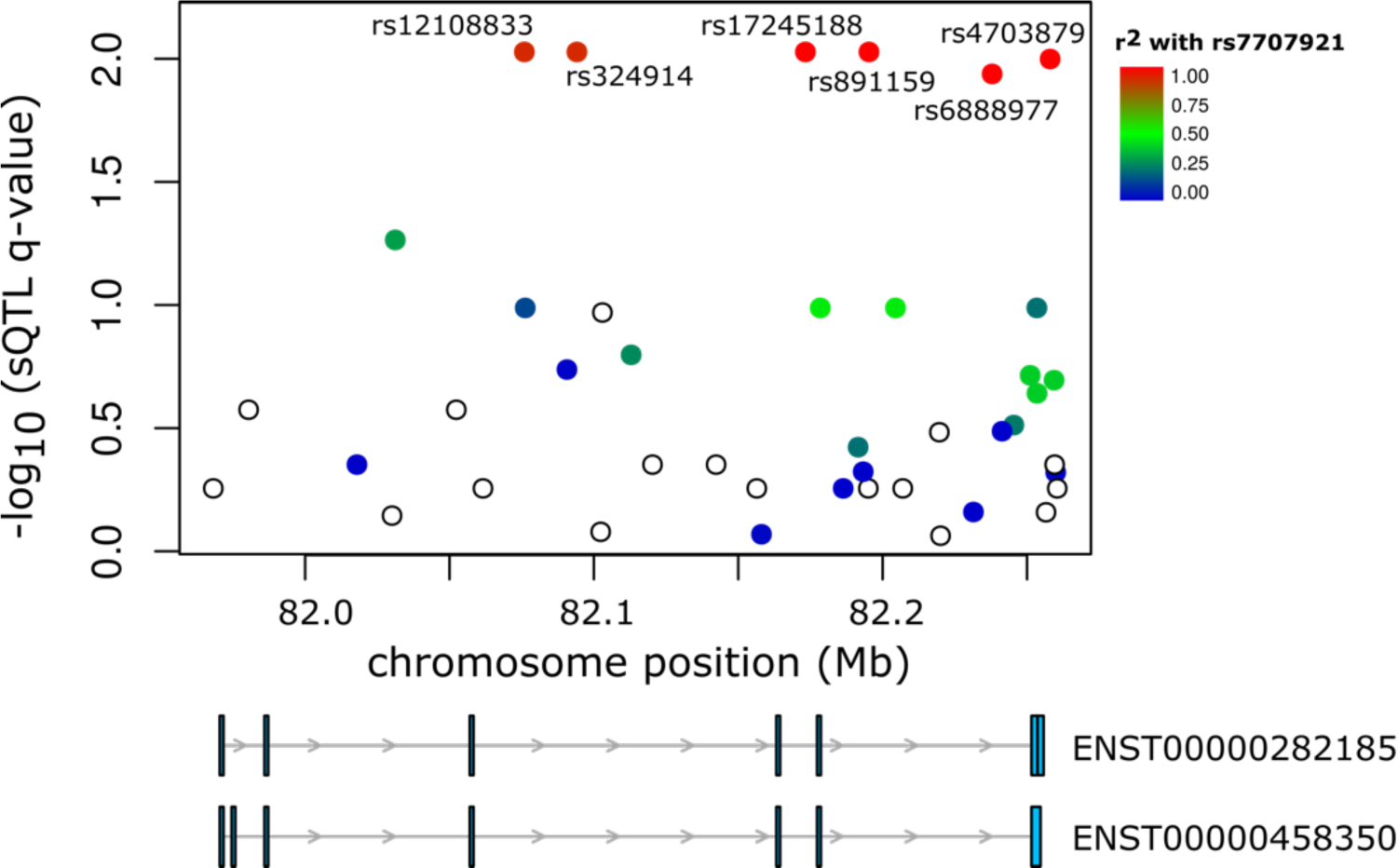
Variants at the 5q14.1 risk locus associated with alternative transcription. Six sQTLs in high LD with rs7707921 were identified for *ATG10*. The -log10(q-value) for the sQTL analysis (y-axis) is shown for the 5q14.1-14.2 region (hg38). Color intensity represents the LD (r^2^) between the analyzed variants and the GWAS lead SNP rs7707921. Below are two *ATG10* transcripts whose expression ratios are associated with the sQTLs.

### Risk model for 5q14.1 links higher expression of *ATG10* and *ATP6AP1L* with protection against BC

Haplotype analysis of the samples included herein revealed two common haplotypes: one harboring the major alleles of all proposed risk-rSNPs and the GWAS lead SNP rs7707921 (frequency of 71.1%) and another with the corresponding minor alleles (frequency of 21.9%) (Figure S21). The proposed risk-rSNPs are among the most significant eQTLs for the two genes: rs111549985 for *ATG10* and rs6880209 for *ATP6AP1L* (Figure S22) (Lonsdale et al., 2013). Therefore, the most common haplotype is associated with an increased risk for BC and lower expression of *ATG10* and *ATP6AP1L* (Figure S21).

Our proposed model for risk at 5q14.1 (Figure 8) establishes that the minor alleles of rs111549985, rs226198, and rs6880209 confer protection against BC by (1) increasing the binding of POL2 II to the promoter of *ATG10* (driven by rs111549985), (2) the binding of POL2 to the shared promoter of *RPS23/ATP6AP1L* (driven by rs6880209), and (3) the binding of cMYC and MAX to a regulatory region (possible enhancer) (driven by rs226198), increasing the expression of *ATG10* and *ATP6AP1L.* These results reveal a complex regulatory landscape at the 5q14.1-14.2 locus, with multiple independent causal variants.

**Figure 8.**
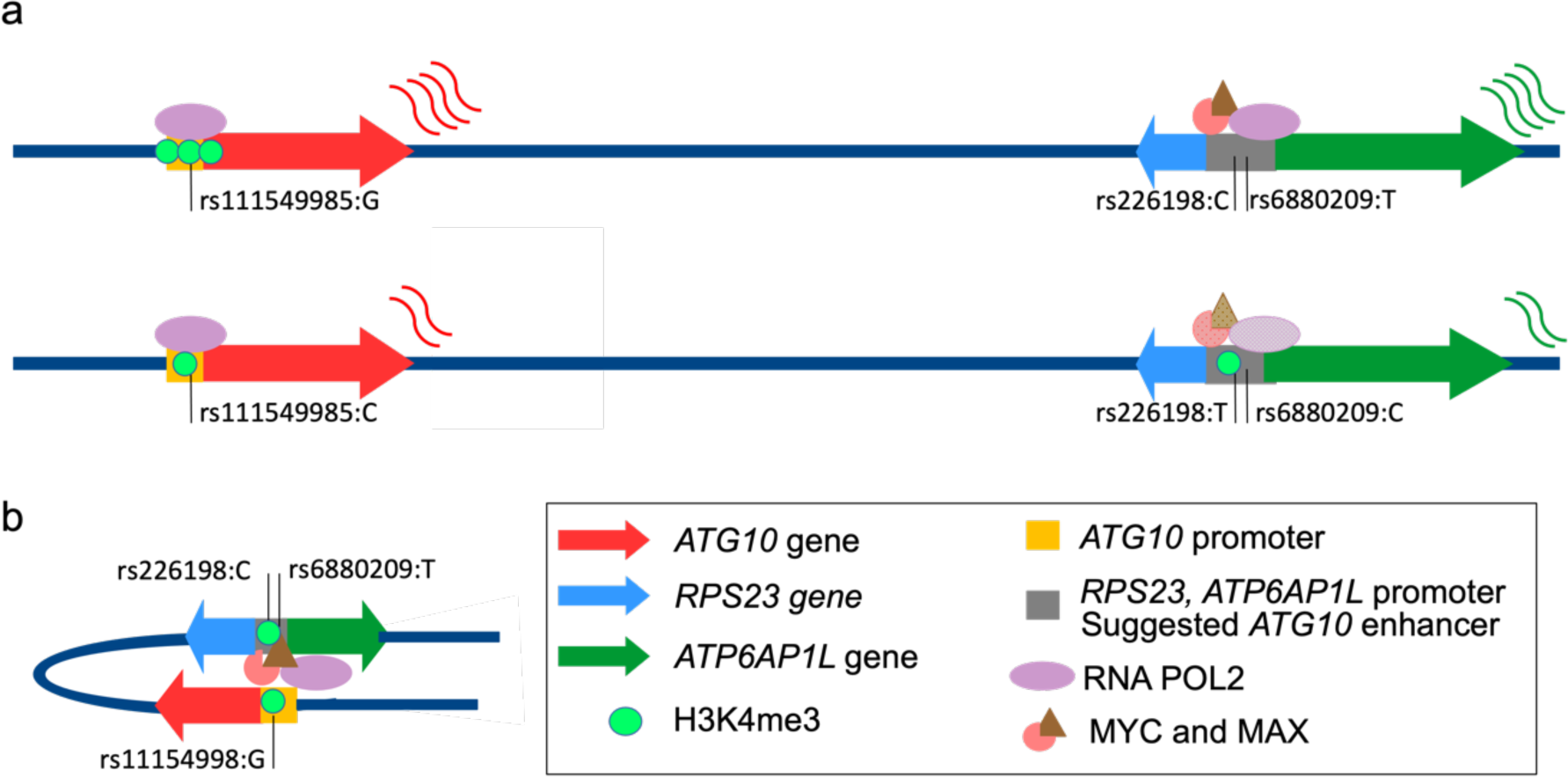
Complex risk regulatory landscape of the 5q14.1 locus. a) Levels of expression of *ATG10* and *ATP6AP1L* genes differ between the haplotypes containing either the minor alleles of rs111549985, rs226198, and rs6880209 (above) or the major ones (below). Colored arrows indicate the direction of transcription of the individual genes, the saturation of the corresponding colors indicates the strength of protein binding, the number of green circles indicates the level of H3K4me3, and the colored curvy lines indicate the relative levels of transcript produced. b) Schematic representation of the proposed model for the positive correlation between *ATG10* and *ATP6AP1L* via a shared regulatory region.

## DISCUSSION

Here, we present the first genome-wide map of differentially expressed allelic genes (daeGenes) in normal breast tissue and their genetic determinants (daeQTLs). We found widespread differential allelic expression (DAE) across the genome and identified daeQTLs for 26% of daeGenes. By intersecting this map with GWAS data, we identified risk-daeQTLs and target genes for 93 BC risk loci. Next, we retrieved epigenetic regulatory annotations on all candidate risk-rSNPs (risk-daeQTLs and their proxies in high LD) to prioritize variants with regulatory potential for further functional analysis. We identified 406 variants with strong regulatory potential annotated to 55 different chromosomal bands and candidates for regulating the expression levels of 96 genes. Our results represent a practical and valuable resource for prioritizing loci for follow-up GWASs. As a proof of concept, we functionally characterized the 5q14.1-14.2 BC risk locus in depth and proposed four causal regulatory variants targeting the genes *ATG10* and *ATP6AP1L* acting via multiple allele-specific mechanisms. Our results suggest a complex regulatory landscape underlying BC etiology.

We show that cis-acting variants regulate the expression of 65% of genes in normal breast tissue, with some genes displaying extreme allelic differences of up to 32-fold. Notably, we identified a novel gene with monoallelic expression, *ARCN1*, which warrants further inspection to confirm its imprinting status. An enrichment of daeSNPs at intergenic and intronic regions, as well as noncoding transcripts, noncoding genes, and pseudogenes, concurs with previous reports of predominant allelic imbalances of expression at gene-depleted regions and genes under fewer evolutionary constraints (Campbell et al., 2008; Tung et al., 2009).

To overcome the lack of phasing information, we applied two different tests in the daeQTL mapping, according to the AE ratio distribution, which led to the identification of 54357 variants associated with AE ratios for 6761 genes, both coding and noncoding for proteins. The stringent statistical correction and the use of distance as a covariate in the second mapping approach increased its confidence level but limited the statistical power to identify regulatory variants in lower LD with the daeSNP or located more distally.

We found evidence of expression regulation by cis-acting variants for most reported GWAS loci and believe that alternative mechanisms are at play in the remainder. We identified risk-daeQTLs at 93 different loci, including 72 loci with novel candidate risk target genes (including *NEK10* at 3p24.1 and *ZBED6* and *ZC3H11A* at 1q32.1). Moreover, the initial daeQTL map in normal breast tissue can be further mined whenever new risk variants are identified through GWAS. These results offer a resource platform for functional studies of causal variants and target genes and can help uncover the role of cis-regulatory variation in BC risk.

Finally, we conducted an *in silico* functional analysis of the 5q14.1-14.2 BC risk locus and identified three strong candidate causal variants: rs111549985, rs226198, and rs6880209. We predict that these variants functionally impact TF binding, chromatin state, and gene expression levels of *ATG10* and *ATP6AP1L*. A similar involvement of diverse regulatory mechanisms has been suggested previously for other BC risk loci (Cox et al., 2011; Fachal et al., 2020; Maia et al., 2012). Both A*TG10* (involved in autophagy) and the *ATP6AP1L* pseudogene have been suggested to have roles in cancer (Jo et al., 2017, 2012; Ma et al., 2021; Wang et al., 2014). A variant at *ATG10* (rs7313473) was previously associated with BC risk by regulating promoter activity, and *ATG10* was suggested to act as a tumor suppressor gene in breast tissue (Guo et al., 2018). For *ATP6AP1L*, another variant (rs10514231) was reported to lead to *ATP6AP1L* downregulation by decreasing the binding affinity of TCF7L2 in an intronic regulatory region (Ma et al., 2021). Although we did not find supporting evidence for the same variants, our results show an indirect association between the lower expression of *ATG10* and *ATP6AP1L* and BC risk, suggesting that the downregulation of these two genes may contribute to tumorigenesis.

The advantages of our analysis compared to previous reports of AE in normal breast and tumor tissue (Aguet et al., 2017; Gao et al., 2012; Przytycki and Singh, 2020; Zhang et al., 2009) include using the most significant number of normal breast tissue samples, the genome-wide approach, and the mapping of candidate regulatory variants. We found a similar frequency of daeSNPs to previous reports in other tissues/cell lines but a higher frequency of daeGenes (Ge et al., 2009; Ma et al., 2018; Przytycki and Singh, 2020; Romanel et al., 2015). This higher frequency of daeGenes could be due to our ability to identify genes regulated by common cis-acting variants with weak to large effect sizes (Aguet et al., 2017), a consequence of the imposed conditions to call DAE (allelic change difference of 1.5-fold and the minimum number of heterozygotes). Additionally, we did not integrate the AE ratios of multiple daeSNPs in the same gene due to the absence of phase data and to maximize the information withdrawn from daeSNPs that might be located in different LD blocks. The complex regulatory landscape we identified at the 5q14.1 locus, with multiple cis-acting variants located in the same haplotypes and AE likely resulting from the sum of the effects of each variant, supports this analysis approach. Furthermore, as we propose, a global measure of the AE imbalance at each gene would impair the mapping of daeQTLs at individual daeSNPs and restrict the analysis to genes with multiple daeSNPs. Finally, besides the more commonly studied protein-coding genes, we analyzed noncoding genes and pseudogenes, such as *ATP6AP1L*.

Our results confirm the advantage of using DAE analysis to detect the effect of rSNPs compared to eQTL analysis, as shown by the higher number of daeGenes than eGenes among gwasGenes (Adoue et al., 2014; Almlöf et al., 2012; Pastinen and Hudson, 2004). As a minority of gwasGenes were exclusively eGenes, we believe that DAE and eQTL analyses are complementary and should be used in parallel when possible.

Our use of microarray data could be seen as a limitation compared to RNA-seq data, which have more extensive transcriptome coverage and high quantification accuracy for more extreme allelic imbalances. However, microarrays are a widely used and precise technology for measuring AE (Gao et al., 2012; Ge et al., 2009; Liu et al., 2012), as we confirmed with our validated monoallelic expression of known imprinted genes and with independent PCR analysis. The only RNA-seq dataset with normal breast tissue publicly available is from the GTEx project. However, our approach presents several advantages: 1) we processed and hybridized the DNA and RNA samples in parallel to minimize technical issues, 2) we used total RNA, which includes coding/noncoding genes and spliced/unspliced transcripts, and 3) we showed that the range of gene expression levels of daeGenes was comparable to that of the eGenes from the GTEx dataset. The following steps will be to carry out matched RNA-seq and DNA-seq to combine all the advantages mentioned above and expand the discovery of daeGenes and rSNPs.

Here, we provide a genome-wide list of variants with strong regulating potential for normal breast tissue, a valuable resource for researchers prioritizing GWAS results for functional characterization and those interested in other BC-related traits. The extensive characterization of the regulatory landscape at the 5q14.1 BC risk locus identified candidate causal variants and revealed the multiple mechanisms involved. Further studies of this locus will elucidate the mechanisms involved and the relative contributions of each variant and target gene to the genetic risk. Overall, our results reinforce the importance of cis-regulatory variation as a major player in BC susceptibility and the power of identifying these variants in the disease’s tissue of origin - normal breast tissue. They also show that multiple causal variants may co-occur and act via independent cis-regulatory mechanisms at BC risk loci, supporting a broader approach to functional studies.

## Data Availability

All data produced are available online at https://github.com/maialab/DaeBreastMicroarrays.

## DECLARATIONS

### Declaration of interests

The authors declare no competing interests.

## Acknowledgments

This work was supported by national Portuguese funding through FCT – Fundação para a Ciência e a Tecnologia and CRESC ALGARVE 2020, institutional support CBMR-UID/BIM/04773/2013, POCI “01-0145-” EDER-022184 “GenomePT”, the contract DL 57/2016/CP1361/CT0042 (J.M.X.) and individual postdoctoral fellowship SFRH/BPD/99502/2014 (J.M.X.). The research leading to these results has received funding from the People Programme (Marie Curie Actions) of the European Union’s Seventh Framework Programme FP7/2007-2013/under REA grant agreement no. 303745 (A.T.M.), a Maratona da Saúde Award (A.T.M.) and a BCRF project Grant.

The authors would also like to thank Dr Nuno Barbosa-Morais at IMM for the excellent scientific discussions and the support given by the Unidade de Apoio à Investigação (UAIC) at Universidade do Algarve (UAlg), particularly Mr. Vitor Morais, and the Informatics Services of UAlg.

## Authors’ Contributions

Conceptualization: Ponder BAJ, Maia AT

Investigation: de Almeida BP, Rocha CL, Rosli N, O’Reilly M, Maia AT

Formal Analysis: Xavier JM, Magno Ramiro, de Almeida BP, Jacinta-Fernandes A, Russell R, Samarajiwa S, Dunning M, António M. Maia, Maia AT

Funding acquisition: Ponder BAJ, Maia AT

Project administration: Maia AT

Writing – original draft: Xavier JM, Maia AT

Writing – review & editing: All authors

## Web Resources

The datasets supporting the conclusions of this article are available in the following repositories: GWAS Catalog [https://www.ebi.ac.uk/gwas], Ensembl database (annotated to GRCh38.p13) [https://www.ensembl.org], GTEx Project (v7 and v8) [https://www.gtexportal.org], microarray data from GEO [https://www.ncbi.nlm.nih.gov/geo/] under accession number GSE35023.

## Data and code availability

The code for this article is available here: https://github.com/maialab/DaeBreastMicroarrays

## Notes

### Competing Interest Statement

The authors have declared no competing interest.

### Author Declarations

Seventy-six samples of normal breast tissue were collected from women submitted to a reduction mastectomy, for reasons unrelated to cancer, at Addenbrooke's Hospital, Cambridge, United Kingdom. Samples were collected with approval from Addenbrooke's Hospital Local Research Ethics Committee (REC reference 06/Q0108/221).

### Summary of Updates

This version of the manuscript has been revised to update the following: - use of a new genome-wide DAE analysis method applying an equal or Given Proportions test and subsequent correction for multiple testing; - use of a new daeQTL mapping method using one-sample Wilcox or a two-sample Wilcox test based on the pattern of the allelic expression ratio distribution displayed at each daeSNP, and subsequent correction for multiple testing; - inclusion of a new section assessing the risk-daeQTLs identified for their cis-acting regulatory potential.

