## Supplementary material for "Mapping of cis-regulatory variants by differential allelic expression analysis identifies candidate causal variants and target genes of 41 breast cancer risk loci": Tables

Joana M. Xavier et al.

**Table 1. Summary of the genome-wide breast tissue allelic expression analysis results**

| Set of SNPs | n | Ensembl Gene IDs |
|---|---|---|
| *All aeSNPs* | 91467 | 21527 |
| *maeSNPs* | 85 | 44 |
| *daeSNPs* | 26,266 | 13,689 |

### Table 2. Loci with candidate risk rSNPs and novel suggested target genes.

| Chr_band | GWAS variant | GWAS nearest gene | Candidate causal variants | daeQTL gene | Regulatory Feature |
|---|---|---|---|---|---|
| **A. Loci with novel suggested target genes** | | | | | |
| 1p36.23 | rs225132 | ERRFI1-DT | rs12757968 | ERRFI1-DT | Active enhancer |
| 1q22 | rs10796944, rs7524950 | ASH1L, PKLR | rs1046188 | FDPS, RUSC1-AS1 | Active promoter and active enhancer |
| 1q22 | rs348196, rs10796944, rs12091730, rs7524950 | DAP3, ASH1L, MSTO1, DAP3P1, PKLR | rs2048431 | FDPS, RUSC1-AS1 | Active promoter and active enhancer |
| 1q22 | rs348196 | DAP3 | rs3841838 | ARHGEF2, RIT1 | Active promoter and active enhancer |
| 1p23.2 | rs67073037 | WDR43 | rs4407214 | WDR43 | Active promoter and active enhancer |
| 4p12 | rs199501877 | NIPAL1, TXK | rs98270 | NIPAL1 | Active promoter and active enhancer |
| 5q11.1 | rs145106188 | EMB | rs4865698 | EMB | Active enhancer |
| 5q11.1 | rs145106188 | EMB | rs4865699 | EMB | Active enhancer |
| 5q11.1 | rs145106188 | EMB | rs28528780 | EMB | Active promoter and active enhancer |
| 5q31.1 | rs6860806 | SLC22A4 | rs1979981 | SLC22A4, MIR3936HG, SLC22A5 | Active promoter and active enhancer |
| 5q31.1 | rs6860806 | SLC22A4 | rs162887 | SLC22A4, MIR3936HG, SLC22A5 | Active promoter and active enhancer |
| 5q31.1 | rs6860806 | SLC22A4 | rs460089 | SLC22A4, MIR3936HG, SLC22A5 | Active promoter and active enhancer |
| 5q31.1 | rs6860806 | SLC22A4 | rs460271 | SLC22A4, MIR3936HG, SLC22A5 | Active promoter and active enhancer |
| 5q31.1 | rs6860806 | SLC22A4 | rs367805 | SLC22A4, MIR3936HG, SLC22A5 | Active enhancer |
| 5q31.1 | rs6860806 | SLC22A4 | rs2631369 | SLC22A4, MIR3936HG, SLC22A5 | Active promoter and active enhancer |
| 5q31.1 | rs6860806 | SLC22A4 | rs2631368 | SLC22A4, MIR3936HG, SLC22A5 | Active promoter and active enhancer |
| 5q31.1 | rs736801 | ENSG00000283782 | rs2070721 | ENSG00000283782 | Active promoter and active enhancer |
| 5q31.1 | rs736801 | ENSG00000283782 | rs2548998 | ENSG00000283782 | Active promoter and active enhancer |
| 6p22.2 | rs17598658, rs13195401 | H2BC6, BTN2A1 | rs9467701 | BTN3A2 | Active promoter and active enhancer |
| 6p22.2 | rs17598658, rs13195401 | H2BC6, BTN2A1 | rs6923139 | BTN3A2 | Active promoter and active enhancer |
| 6p22.2 | rs17598658, rs13195401 | H2BC6, BTN2A1 | rs6903015 | BTN3A2 | Active promoter and active enhancer |
| 6p22.2 | rs17598658, rs13195401 | H2BC6, BTN2A1 | rs68112369 | BTN3A2 | Active promoter |
| 6p22.2 | rs17598658, rs13195401 | H2BC6, BTN2A1 | rs66827971 | BTN3A2 | Active promoter |
| 6p22.2 | rs13195401 | BTN2A1 | rs9379873 | BTN3A2 | Active promoter and active enhancer |
| 6p22.2 | rs71557345, rs13195401 | ENSG00000285571, BTN2A1 | rs36162392 | BTN3A2 | Active promoter and active enhancer |

| | | | | | |
|---|---|---|---|---|---|
| 6p22.1 | rs3094146, rs1611579 | ZNRD1ASP, ENSG00000285799 | rs707910 | HLA-A | Active promoter and active enhancer |
| 6p22.1 | rs3094146, rs1611579 | ZNRD1ASP, ENSG00000285799 | rs415137 | HLA-A | Active promoter and active enhancer |
| 6p22.1 | rs3094146, rs1611579 | ZNRD1ASP, ENSG00000285799 | rs438610 | HLA-A | Active promoter and active enhancer |
| 6p22.1 | rs3094054, rs3094146, rs3132615 | UBQLN1P1, ZNRD1ASP, ENSG00000288805 | rs2188100 | HCG17, HLA-L | Active promoter and active enhancer |
| 6p22.1 | rs3129984, rs3132610, rs9262142, rs3132615 | ENSG00000288805, HCG20, ABCF1, PPP1R18 | rs9262142 | C6orf136 | Active promoter and active enhancer |
| 7q21.3 | rs847577 | LMTK2 | rs1874343 | TECPR1 | Active promoter and active enhancer |
| 9q31.1 | rs4742903, rs718857 | SMC2 | rs3818625 | SMC2 | Active promoter |
| 9q31.1 | rs4742903, rs718857 | SMC2 | rs3818626 | SMC2 | Active promoter |
| 9q31.1 | rs4742903, rs718857 | SMC2 | rs10820599 | SMC2 | Active promoter |
| 9q31.1 | rs4742903, rs718857 | SMC2 | rs10820600 | SMC2 | Active promoter |
| 9q31.1 | rs4742903, rs718857 | SMC2 | rs4742903 | SMC2 | Active promoter |
| 10p12.31 | rs7072776, rs10828247, rs11012730, rs10828249, rs7098100 | MLLT10, SKIDA1 | rs10828247 | MLLT10 | Active promoter |
| 10p12.1 | rs7918232 | ANKRD26 | rs7907988 | YME1L1 | Active promoter and active enhancer |
| 10q21.2 | rs10822013 | ENSG00000285837, ZNF365 | rs10822013 | ENSG00000285837 | Active enhancer |
| 11p11.2 | rs11039183 | MADD | rs7947450 | SPI1 | Active promoter |
| 11q13.1 | rs617791 | DRAP1, TSGA10IP | rs14157 | SART1 | Active promoter |
| 11q13.1 | rs617791 | DRAP1, TSGA10IP | rs12794370 | SART1 | Active promoter and active enhancer |
| 11q13.2 | rs1783730 | TMEM151A | rs33981819 | ENSG00000254458 | Active promoter and active enhancer |
| 11q13.2 | rs11344495, rs55908905 | CTSF, SPTBN2 | rs11110 | DPP3-DT, B4GAT1-DT | Active promoter and active enhancer |
| 11q13.2 | rs11344495, rs55908905 | CTSF, SPTBN2 | rs55853079 | DPP3-DT, B4GAT1-DT | Active promoter and active enhancer |
| 11q22.3 | rs11374964 | POGLUT3 | rs228589 | NPAT | Active promoter |
| 11q22.3 | rs11374964 | POGLUT3 | rs189037 | NPAT | Active promoter |
| 11q23.1 | rs505372 | SIK2 | rs541198 | NA | Active promoter and active enhancer |
| 11q23.1 | rs505372 | SIK2 | rs7107213 | CRYAB | Active promoter and active enhancer |
| 14q13.2 | rs58327846 | PRORP | rs1056879 | PRORP | Active promoter and active enhancer |
| 14q13.2 | rs58327846 | PRORP | rs2236167 | PRORP | Active promoter and active enhancer |
| 14q32.33 | rs60226654 | COA8 | rs3759586 | KLC1, XRCC3 | Active enhancer |

| | | | | | |
|---|---|---|---|---|---|
| 15q21.1 | rs1876206 | FBN1 | rs8029993 | FBN1 | Active enhancer |
| 15q24.2 | rs60381548, rs8027365 | SIN3A, PTPN9 | rs11637068 | MAN2C1, PTPN9 | Active promoter and active enhancer |
| 15q24.2 | rs60381548, rs8027365 | SIN3A, PTPN9 | rs75219778 | MAN2C1, PTPN9 | Active promoter |
| 15q24.2 | rs60381548, rs8027365 | SIN3A, PTPN9 | rs62027209 | MAN2C1, PTPN9 | Active promoter and active enhancer |
| 15q26.1 | rs77554484 | PRC1, ENSG00000284946 | rs867468 | PRC1, PRC1-AS1, ENSG00000284946 | Active promoter and active enhancer |
| 15q26.1 | rs77554484 | PRC1, ENSG00000284946 | rs2001216 | PRC1, PRC1-AS1, ENSG00000284946 | Active promoter and active enhancer |
| 15q26.1 | rs77554484 | PRC1, ENSG00000284946 | rs12905855 | PRC1, PRC1-AS1, ENSG00000284946 | Active promoter and active enhancer |
| 16q12.2 | rs17817449, rs62033406, rs7193144, rs62048402 | FTO | rs9940128 | FTO | Active enhancer |
| 16q12.2 | rs17817449, rs62033406, rs7193144, rs62048402 | FTO | rs11642015 | FTO | Active enhancer |
| 16q12.2 | rs17817449, rs62033406, rs7193144, rs62048402 | FTO | rs17817497 | FTO | Active enhancer |
| 16q12.2 | rs17817449, rs62033406, rs7193144, rs62048402 | FTO | rs8050136 | FTO | Active enhancer |
| 16q13 | rs2303282, rs2432539 | BBS2, AMFR | rs2440467 | AMFR | Active enhancer |
| 17q21.31 | rs2732699 | ARL17B | rs76594404 | CRHR1, KANSL1, LINC02210, LINC02210-CRHR1, MAPT, MAPT-AS1, ARL17B | Active promoter and active enhancer |
| 17q21.31 | rs2732699 | ARL17B | rs80233201 | CRHR1, KANSL1, LINC02210, LINC02210-CRHR1, MAPT, MAPT-AS1, ARL17B | Active promoter and active enhancer |
| 17q21.31 | rs2732699 | ARL17B | rs62056778 | CRHR1, KANSL1, LINC02210, LINC02210-CRHR1, MAPT, MAPT-AS1, ARL17B | Active promoter and active enhancer |
| 17q21.31 | rs2732699 | ARL17B | rs11575895 | CRHR1, KANSL1, LINC02210, LINC02210-CRHR1, MAPT, MAPT-AS1, ARL17B | Active promoter |
| 17q21.31 | rs2732699 | ARL17B | rs62056779 | CRHR1, KANSL1, LINC02210, LINC02210-CRHR1, MAPT, MAPT-AS1, ARL17B | Active promoter |
| 17q21.31 | rs2732699 | ARL17B | rs74548327 | CRHR1, KANSL1, LINC02210, LINC02210-CRHR1, MAPT, MAPT-AS1, ARL17B | Active promoter |
| 17q21.31 | rs2732699 | ARL17B | rs111972148 | CRHR1, KANSL1, LINC02210, LINC02210-CRHR1, MAPT, MAPT-AS1, ARL17B | Active promoter |

| | | | | | |
|---|---|---|---|---|---|
| 17q21.31 | rs2732699 | ARL17B | rs242561 | CRHR1, KANSL1, LINC02210, LINC02210-CRHR1, MAPT, MAPT-AS1, ARL17B | Active promoter |
| 17q21.31 | rs2732699 | ARL17B | rs2316951 | CRHR1, KANSL1, LINC02210, LINC02210-CRHR1, MAPT, MAPT-AS1, ARL17B | Active enhancer |
| 17q21.31 | rs2732699 | ARL17B | rs11079733 | CRHR1, KANSL1, LINC02210, LINC02210-CRHR1, MAPT, MAPT-AS1, ARL17B | Active promoter |
| 17q21.31 | rs2732699 | ARL17B | rs2696633 | CRHR1, KANSL1, LINC02210, LINC02210-CRHR1, MAPT, MAPT-AS1, ARL17B | Active promoter |
| 17q21.31 | rs2732699 | ARL17B | rs143625699 | ARL17B, KANSL1 | Active promoter |
| 17q21.31 | rs2732699 | ARL17B | rs113417378 | CRHR1, KANSL1, LINC02210, LINC02210-CRHR1, MAPT, MAPT-AS1, ARL17B | Active promoter |
| 17q21.31 | rs2732699 | ARL17B | rs143191191 | CRHR1, KANSL1, LINC02210, LINC02210-CRHR1, MAPT, MAPT-AS1, ARL17B | Active enhancer |
| 22q13.31 | rs28512361 | ENSG00000235091 | rs134847 | ATXN10 | Active promoter |
| 22q13.31 | rs28512361 | ENSG00000235091 | rs2071872 | ATXN10 | Active promoter and active enhancer |
| **B. Loci with previously suggested target genes** | | | | | |
| 1p34.1 | rs12077974 | MAST2 | rs6697821 | NASP#, IPP# | Active promoter and active enhancer |
| 1p34.1 | rs12077974, rs1707302 | MAST2, PIK3R3 | rs1707303 | NASP#, IPP#, PIK3R3 | Active promoter and active enhancer |
| 1p34.1 | rs12077974 | MAST2 | rs1707302 | NASP#, IPP#, PIK3R3 | Active enhancer |
| 2q33.1 | rs10931936, rs1035142, rs1830298, rs700635, rs3769821 | CASP8 | rs3769823 | CASP8 | Active promoter and active enhancer |
| 3q12.1 | rs9837602, rs9833888, rs9289981 | CMSS1, FILIP1L | rs793463 | CMSS1#, FILIP1L | Active enhancer |
| 3q12.1 | rs9837602, rs9833888 | CMSS1, FILIP1L | rs28714363 | CMSS1#, FILIP1L | Active enhancer |
| 5q14.1 | rs7707921, rs111549985, rs2407064, rs146817970, rs2407156 | ATG10, ATP6AP1L | rs111549985 | ATG10, ATP6AP1L | Active promoter and active enhancer |
| 5q14.1 | rs7707921, rs2407064, rs146817970, rs2407156 | ATG10 | rs226198 | ATG10, ATP6AP1L | Active promoter and active enhancer |
| 5q14.1 | rs7707921, rs2407064, rs146817970, rs2407156 | ATG10 | rs6880209 | ATG10, ATP6AP1L | Active promoter and active enhancer |
| 5q14.1 | rs7707921, rs2407064, rs146817970, rs2407156 | ATG10 | rs11325430 | ATG10, ATP6AP1L | Active promoter and active enhancer |

| | | | | | |
|---|---|---|---|---|---|
| 5q14.1 | rs7707921, rs2407064, rs146817970 | ATG10 | rs17247678 | ATG10, ATP6AP1L | Active enhancer |
| 7q21.2 | rs6964587, rs10644111, rs35417517 | AKAP9, LRRD1, CYP51A1-AS1 | rs1011372 | AKAP9 | Active promoter and active enhancer |
| 7q21.2 | rs35522438, rs35417517, rs10644111, rs6964587 | AKAP9, LRRD1, CYP51A1-AS1, KRIT1 | rs5885795 | AKAP9 | Active promoter and active enhancer |
| 7q21.2 | rs35522438, rs35417517, rs10644111, rs6964587 | AKAP9, LRRD1, CYP51A1-AS1, KRIT1 | rs4727266 | AKAP9 | Active promoter and active enhancer |
| 7q21.2 | rs35522438, rs35417517, rs10644111, rs6964587 | AKAP9, LRRD1, CYP51A1-AS1, KRIT1 | rs4727267 | AKAP9 | Active promoter and active enhancer |
| 7q21.2 | rs35522438, rs35417517, rs10644111, rs6964587 | AKAP9, LRRD1, CYP51A1-AS1, KRIT1 | rs6465339 | AKAP9 | Active promoter and active enhancer |
| 7q21.2 | rs35522438, rs35417517, rs10644111, rs6964587 | AKAP9, LRRD1, CYP51A1-AS1, KRIT1 | rs4279 | AKAP9 | Active promoter and active enhancer |
| 7q21.2 | rs35522438, rs35417517, rs10644111, rs6964587 | AKAP9, LRRD1, CYP51A1-AS1, KRIT1 | rs12704637 | AKAP9 | Active promoter and active enhancer |
| 11p15.5 | rs6597981 | PIDD1 | rs7942564 | GATD1 | Active promoter and active enhancer |
| 11p15.5 | rs6597981 | PIDD1 | rs7948070 | GATD1 | Active promoter and active enhancer |
| 11q13.1 | rs3903072 | SNX32, OVOL1 | rs4621 | CFL1, SNX32# | Active promoter and active enhancer |
| 11q13.1 | rs3903072 | SNX32, OVOL1 | rs7125986 | CFL1, SNX32# | Active promoter |
| 11q13.1 | rs3903072 | SNX32, OVOL1 | rs7947929 | CFL1, SNX32# | Active promoter |
| 11q13.1 | rs3903072 | SNX32, OVOL1 | rs7947741 | CFL1, SNX32# | Active promoter |
| 11q13.1 | rs3903072 | SNX32, OVOL1 | rs13817 | CFL1, SNX32# | Active promoter and active enhancer |
| 19p13.11 | rs8170 | BABAM1 | rs3745187 | ABHD8 | Active promoter |
| 19p13.11 | rs2965183, rs2304098 | GATAD2A, YJEFN3 | rs3934667 | MAU2 | Active promoter |
| 19p13.11 | rs2965183, rs2304098 | GATAD2A, YJEFN3 | rs2916068 | MAU2 | Active promoter |
| 19p13.11 | rs2965183, rs2304098 | GATAD2A, YJEFN3 | rs80007081 | YJEFN3#, CILP2#, ENSG00000258674# | Active promoter and active enhancer |
| 19p13.11 | rs2965183, rs2304098 | GATAD2A, YJEFN3 | rs17684164 | YJEFN3#, CILP2#, ENSG00000258674# | Active promoter and active enhancer |
| 19p13.11 | rs2965183, rs2304098 | GATAD2A, YJEFN3 | rs77254326 | YJEFN3#, CILP2#, ENSG00000258674# | Active promoter and active enhancer |
| **C. Loci for which no clear target gene can be pointed** | | | | | |
| 6p22.1 | rs9257408 | KRT18P1 | rs209174 | NA | Active promoter and active enhancer |
| 6p22.1 | rs9257408 | KRT18P1 | rs209173 | NA | Active promoter and active enhancer |

| | | | | | |
|---|---|---|---|---|---|
| 6p22.1 | rs9257408 | KRT18P1 | rs3135315 | NA | Active promoter and active enhancer |
| 6p22.1 | rs9257408 | KRT18P1 | rs184093 | NA | Active promoter and active enhancer |
| 6p22.1 | rs9257408 | KRT18P1 | rs209138 | NA | Active promoter |
| 6p22.1 | rs9257408 | KRT18P1 | rs3131102 | NA | Active promoter |
| 17q23.1 | rs61495451 | VMP1 | rs3803863 | chr17:59852174 | Active promoter |
| 17q23.1 | rs61495451 | VMP1 | rs2333562 | chr17:59852174 | Active promoter and active enhancer |
| 17q23.1 | rs61495451 | VMP1 | rs138148328 | chr17:59852174 | Active enhancer |
| 19p13.11 | rs4808801, rs172032, rs7258465 | ELL, SSBP4 | rs28375303 | chr19:18416447 | Active promoter |
| 19p13.11 | rs4808801, rs172032, rs7258465 | ELL, SSBP4 | rs271621 | chr19:18416447 | Active promoter and active enhancer |

Legend: * Reported as rs116095464 in the original GWAS

[#] Linkage disequilibrium (LD) values $r^2$ between the daeQTL and the GWAS risk variant in the European population

[§] Gene not expressed in breast mammary tissue or without expression information in GTEx
