## Supplemental Tables for "Mapping of cis-regulatory variants by differential allelic expression analysis identifies candidate causal variants and target genes of 41 breast cancer risk loci"

Joana M. Xavier *et al*.

**Table S1:** SNPs associated with BC risk retrieved from GWAS Catalog using a significance threshold for association signals of p-value ≤1.0E-05.

**Access Link: Xavier_et_al_2023_TableS1**

**Table S2: Previously reported target genes of cis-acting regulatory variation in post-GWAS studies for breast cancer.** Information includes whether daeSNPs were identified for each gene (daeGene) and whether they were reported by GTEx (v7) as having significant association with eQTLs variants (eGenes) in breast mammary tissue.

**Access Link: Xavier_et_al_2023_TableS2**

**Table S3: Genomic annotation of aeSNPs.** Annotations were retrieved according to the Ensembl database (GRCh38.p13).

**Access Link: Xavier_et_al_2023_TableS3**

**Table S4: Annotation of maeSNPs identified (SNPs displaying mono-allelic expression) in normal breast tissue.** Local gene(s) corresponds to the gene(s) annotated within 5Kb of the maeSNP.

#Information retrieved from the Geneimprint database: (https://www.geneimprint.com/site/genes-by-species.Homo+sapiens) and from Goovaerts et al. 2018.

**Access Link: Xavier_et_al_2023_TableS4**

**Table S5: Annotation of significant daeQTLs.** wilcoxon_test_type corresponds to the test used for daeQTL mapping.

**Access Link: Xavier_et_al_2023_TableS5**

**Table S6: Genomic annotation and classification of genes harbouring aeSNPs as daeGenes, eGenes, maeGenes, daeGenes with daeQTL mapping and gwasGenes. Legend:** daeGenes - genes which harbour daeSNPs; eGenes - genes associated with an eQTL variant (GTEx v7, breast mammary tissue); maeGenes - genes which harbour maeSNPs; daeGenes with daeQTLs - genes which harbour significant daeQTLs variants; gwasGenes - genes which harbour GWAS hit variants and/or their proxies (using an LD $r^2 \geq 0.4$).

**Access Link: Xavier_et_al_2023_TableS6**

**Table S7: Annotation of risk-daeQTLs identified. Legend:** risk-daeQTLs were defined as daeQTLs with an $r^2 \geq 0.4$ with a BC GWAS hit variant; Mapping Approach corresponds to the approach used in the daeQTL analysis; Pubmed ID - GWAS_SNP original publication; R_squared - LD between the daeQTL and the GWAS_SNP.

**Access Link: Xavier_et_al_2023_TableS7**

**Table S8: List of candidate causal variants located in regulatory regions.**

**Access Link: Xavier_et_al_2023_TableS8**

**Table S9: List of daeQTLs identified with an $0.2 \leq r^2 < 0.4$ with a BC GWAS hit variant.**

Legend: daeGene_hgnc_symbol - daeSNP gene name; daeQTL_gene_hgnc_symbol - daeQTL gene name; Mapping Approach - approach used in the daeQTL analysis; R_squared - LD between the daeQTL and the GWAS_SNP.

**Access Link: Xavier_et_al_2023_TableS9**

**Table S10: Candidate causal variants for ATG10 and ATP6AP1L located in regions annotated as promoter and/or enhancers.** LD (measured by $r^2$) values between the candidate risk rSNPs and the GWAS lead SNP rs7707921, the p-values from the binomial test and the q-values after correction for multiple testing are indicated.

**Access Link: Xavier_et_al_2023_TableS10**

**Table S11: Allele-specific epigenetic modifications and transcription factors binding information at three candidate risk-rSNPs.** Fold-change corresponds to the ratio between the number of counts at the major allele by the number of counts at the minor allele. The significance from the binomial test before (p-value) and after (q-value) correction for multiple testing are indicated.

**Access Link: Xavier_et_al_2023_TableS11**

**Table S12: sQTL analysis for the ATG10 gene using data from GTEx_MammaryTissue, TCGA-BRCA_Primary-Solid-Tumour and TCGA-BRCA_Solid-Tissue-Normal and carried out with sQTLseeker.** Legend: snpId - SNP name; geneId - gene name; F - F score;

nb.groups - the number of genotype groups (2 or 3); md - the maximum difference in relative expression between genotype groups; tr.first/tr.second - the transcript IDs of the two transcripts that change the most (and symmetrically); nb.perms - the number of permutation used for the P-value computation; pvalue - P-values for each pair of gene/SNP testing the association between the genotype and transcript relative expression; R_squared - LD between the SNP and the GWAS lead SNP rs7707921.

**Access Link: Xavier_et_al_2023_TableS12**

**Table S13: CLIP data of bound RBPs at two candidate risk-rSNPs for the *ATG10* gene.**

**Access Link: Xavier_et_al_2023_TableS13**

**Table S14: Allele-specific RNA Binding Proteins (RBP) predictions for two variants located at *ATG10*.** Alleles are indicated in the order of reference/alternative. RBP_map_Ref and RPB_map_Alt indicate the predicted RBP name next to the predicted binding position (the variant is located at position 31) and p-value is an estimate from a normal one-tailed distribution of the Z-score.

**Access Link: Xavier_et_al_2023_TableS14**

Meyer KB, Maia AT, O'Reilly M, Teschendorff AE, Chin SF, Caldas C, Ponder BA.

Allele-specific up-regulation of FGFR2 increases susceptibility to breast cancer. PLoS Biol. 2008 May 6;6(5):e108.

Meyer KB, O'Reilly M, Michailidou K, Carlebur S, Edwards SL, French JD, et al. Fine-scale mapping of the FGFR2 breast cancer risk locus: putative functional variants differentially bind FOXA1 and E2F1. Am J Hum Genet. 2013 Dec 5;93(6):1046-60.

Michailidou K, Hall P, Gonzalez-Neira A, Ghoussaini M, Dennis J, Milne RL, et al. Large-scale genotyping identifies 41 new loci associated with breast cancer risk. Nat Genet. 2013 Apr;45(4):353-61, 361e1-2.

Michailidou K, Beesley J, Lindstrom S, Canisius S, Dennis J, Lush MJ, et al. Genome-wide association analysis of more than 120,000 individuals identifies 15 new susceptibility loci for breast cancer. Nat Genet. 2015 Apr;47(4):373-80.

Michailidou K, Lindström S, Dennis J, Beesley J, Hui S, Kar S, et al. Association analysis identifies 65 new breast cancer risk loci. Nature. 2017 Nov 2;551(7678):92-94.

Milne RL, Kuchenbaecker KB, Michailidou K, Beesley J, Kar S, Lindström S, et al. Identification of ten variants associated with risk of estrogen-receptor-negative breast cancer. Nat Genet. 2017 Dec;49(12):1767-1778.

Murabito JM, Rosenberg CL, Finger D, Kreger BE, Levy D, Splansky GL, Antman K, Hwang SJ. A genome-wide association study of breast and prostate cancer in the NHLBI's Framingham Heart Study. BMC Med Genet. 2007 Sep 19;8 Suppl 1(Suppl 1):S6.

Orr N, Lemnrau A, Cooke R, Fletcher O, Tomczyk K, Jones M, et al. Genome-wide association study identifies a common variant in RAD51B associated with male breast cancer risk. Nat Genet. 2012 Nov;44(11):1182-4.

Orr N, Dudbridge F, Dryden N, Maguire S, Novo D, Perrakis E, et al. Fine-mapping identifies two additional breast cancer susceptibility loci at 9q31.2. Hum Mol Genet. 2015 May 15;24(10):2966-84.

Palomba G, Loi A, Porcu E, Cossu A, Zara I, Budroni M, Dei M, Lai S, Mulas A, Olmeo N, Ionta MT, Atzori F, Cuccuru G, Pitzalis M, Zoledziewska M, Olla N, Lovicu M, Pisano M, Abecasis GR, Uda M, Tanda F, Michailidou K, Easton DF, Chanock SJ, Hoover RN, Hunter DJ, Schlessinger D, Sanna S, Crisponi L, Palmieri G. Genome-wide association study of susceptibility loci for breast cancer in Sardinian population. BMC Cancer 2015 May 10;15:383.

Pande M, Joon A, Brewster AM, Chen WV, Hopper JL, Eng C, et al. Genetic susceptibility markers for a breast-colorectal cancer phenotype: Exploratory results from genome-wide association studies. PLoS One. 2018 Apr 26;13(4):e0196245.

Quigley DA, Fiorito E, Nord S, Van Loo P, Alnæs GG, Fleischer T, et al. The 5p12 breast cancer susceptibility locus affects MRPS30 expression in estrogen-receptor positive tumors. Mol Oncol. 2014 Mar;8(2):273-84.

Rinella ES, Shao Y, Yackowski L, Pramanik S, Oratz R, Schnabel F, Guha S, LeDuc C, Campbell CL, Klugman SD, Terry MB, Senie RT, Andrulis IL, Daly M, John EM, Roses D, Chung WK, Ostrer H. Genetic variants associated with breast cancer risk for Ashkenazi Jewish women with strong family histories but no identifiable BRCA1/2 mutation. Hum Genet. 2013 May;132(5):523-36.

Sehrawat B, Sridharan M, Ghosh S, Robson P, Cass CE, Mackey JR, Greiner R, Damaraju S. Potential novel candidate polymorphisms identified in genome-wide association study for breast cancer susceptibility. Hum Genet. 2011 Oct;130(4):529-37.

Shi J, Zhang Y, Zheng W, Michailidou K, Ghoussaini M, Bolla MK, et al. Fine-scale mapping of 8q24 locus identifies multiple independent risk variants for breast cancer. Int J Cancer. 2016 Sep 15;139(6):1303-1317.

Siddiq A, Couch FJ, Chen GK, Lindström S, Eccles D, Millikan RC, et al. A meta-analysis of genome-wide association studies of breast cancer identifies two novel susceptibility loci at 6q14 and 20q11. Hum Mol Genet. 2012 Dec 15;21(24):5373-84.

Song C, Chen GK, Millikan RC, Ambrosone CB, John EM, Bernstein L, et al. A genome-wide scan for breast cancer risk haplotypes among African American women. PLoS One. 2013;8(2):e57298.

Stacey SN, Manolescu A, Sulem P, Rafnar T, Gudmundsson J, Gudjonsson SA, et al. Common variants on chromosomes 2q35 and 16q12 confer susceptibility to estrogen receptor-positive breast cancer. Nat Genet. 2007 Jul;39(7):865-9.

Stacey SN, Sulem P, Zanon C, Gudjonsson SA, Thorleifsson G, Helgason A, et al. Ancestry-shift refinement mapping of the C6orf97-ESR1 breast cancer susceptibility locus. PLoS Genet. 2010 Jul 22;6(7):e1001029.

Sun Y, Ye C, Guo X, Wen W, Long J, Gao YT, Shu XO, Zheng W, Cai Q. Evaluation of potential regulatory function of breast cancer risk locus at 6q25.1. Carcinogenesis. 2016 Feb;37(2):163-168.

Thomas G, Jacobs KB, Kraft P, Yeager M, Wacholder S, Cox DG, et al. A multistage genome-wide association study in breast cancer identifies two new risk alleles at 1p11.2 and 14q24.1 (RAD51L1). Nat Genet. 2009 May;41(5):579-84.

Turnbull C, Ahmed S, Morrison J, Pernet D, Renwick A, Maranian M, et al. Genome-wide association study identifies five new breast cancer susceptibility loci. Nat Genet. 2010 Jun;42(6):504-7.

Wang Y, He Y, Qin Z, Jiang Y, Jin G, Ma H, Dai J, Chen J, Hu Z, Guan X, Shen H. Evaluation of functional genetic variants at 6q25.1 and risk of breast cancer in a Chinese population. Breast Cancer Res. 2014 Aug 14;16(4):422.

Wynendaele J, Böhnke A, Leucci E, Nielsen SJ, Lambertz I, Hammer S, et al. An illegitimate microRNA target site within the 3' UTR of MDM4 affects ovarian cancer progression and chemosensitivity. Cancer Res. 2010 Dec 1;70(23):9641-9.

Wyszynski A, Hong CC, Lam K, Michailidou K, Lytle C, Yao S, et al. An intergenic risk locus containing an enhancer deletion in 2q35 modulates breast cancer risk by deregulating IGFBP5 expression. Hum Mol Genet. 2016 Sep 1;25(17):3863-3876.

Zeng C, Guo X, Long J, Kuchenbaecker KB, Droit A, Michailidou K, et al. Identification of independent association signals and putative functional variants for breast cancer risk through fine-scale mapping of the 12p11 locus. Breast Cancer Res. 2016 Jun 21;18(1):64.

Zheng W, Long J, Gao YT, Li C, Zheng Y, Xiang YB, Wen W, Levy S, Deming SL, Haines JL, Gu K, Fair AM, Cai Q, Lu W, Shu XO. Genome-wide association study identifies a new breast cancer susceptibility locus at 6q25.1. Nat Genet. 2009 Mar;41(3):324-8.
